# Border quarantine, vaccination and public health measures to mitigate the impact of COVID-19 importations in Australia: a modelling study

**DOI:** 10.1101/2024.04.22.24305704

**Authors:** Michael J Lydeamore, Cameron Zachreson, Eamon Conway, Freya M Shearer, Christopher M Baker, Joshua V Ross, Joel C Miller, James M McCaw, Nicholas Geard, Jodie McVernon, David J Price

## Abstract

We developed a flexible infectious disease model framework that combines a detailed individual-based model of arrival pathways (quarantine model) and an individual-based model of the arrivals environment (community model) to inform border risk assessments. The work was motivated by Australia’s desire to safely increase international arrival volumes, which had been heavily constrained since early 2020 as a result of the COVID-19 pandemic. These analyses supported decisions on quarantine and border policy in the context of the Australian government’s national reopening plan in late 2021.

The quarantine model provides a detailed representation of transmission within quarantine and time-varying infectiousness and test sensitivity within individuals, to characterise the likelihood and infectiousness of breaches from quarantine. The community model subsequently captures the impact these infectious individuals have in the presence of varying vaccination coverage, arrival volumes, public health and social measures (PHSMs) and test-trace-isolate-quarantine system effectiveness in the Australian context.

Our results showed that high vaccination coverage would be required to safely reopen with support from ongoing PHSMs, and quarantine pathways have minimal impact on infection dynamics in the presence of existing local transmission. The modelling pipeline we present can be flexibly adapted to a range of scenarios, providing a useful framework for timely border and quarantine risk assessment.

## Introduction

Border controls and quarantine have played an important role in the COVID-19 response in many countries by reducing the risk of importation, thereby postponing establishment of community transmission [1–3]. The use of strong border measures enabled many island nations in the Western Pacific region including Australia to minimise the number of seeding events, to the extent that public health and social measures (PHSMs) could maintain very low levels of transmission for extended periods throughout 2020 and 2021 until effective vaccines were available. However, border measures impose considerable social and economic costs by limiting personal and business travel, making reopening essential when deemed safe to do so.

In Australia, the initial wave of COVID-19 in 2020 was brought under control by a nationwide lockdown that supported active case finding and contact management, achieving local elimination. This situation was effectively maintained in most of the country over the next two years by stringent caps on international arrivals and mandatory quarantine requirements [4, 5]. Quarantine was managed by states and territories, using hotels and a small number of repurposed accommodation facilities, all of which had to be adapted and operated according to strict protocols to be fit for purpose. Infection breaches were rare but highly visible, requiring active public health management and social measures including lockdowns to constrain onward spread [6]. These disruptive events prompted close attention to mitigation of residual risks in the arrivals system [5].

Incursions associated with travellers were due either to very prolonged viral shedding (if infected in the country of origin) or undetected infections acquired from fellow travellers during the quarantine period. Transmission to quarantine workers was a more frequently identified source of infection importation into the wider community. Vaccination of travellers and workers within the quarantine system prior to the emergence of the Omicron variant was a highly effective strategy to reduce the risk of transmission events within managed facilities, and hence the rate of breaches [7] — though the reduced effectiveness of vaccines against transmission for Omicron would subsequently lessen this impact [8]. Widespread administration of safe and effective vaccines at the population level further reduces the likelihood of effective seeding and the health and societal impacts of infection, easing pressure on border controls as the front line of population protection.

In August 2021, the Australian government proposed the ‘National Plan to transition Australia’s National COVID-19 Response’ (herein, ‘national reopening plan’) in which national vaccination coverage targets were explicitly tied to easing of restrictions on international travel and the end of lockdowns as an infection control strategy [9, 10]. At this time, the Delta variant of SARS-CoV-2 was circulating globally, with varying epidemiological characteristics across Australian states and territories ranging from jurisdictions that were largely free of COVID-19 to those with ongoing community transmission — correspondingly, jurisdictions had variable vaccination coverage and public health and social measures in place. While vaccine uptake was increasing across Australia, it was unclear what level of ongoing border controls would be required to limit the risk of sustained community outbreaks. Quarantine pathways had to be reconsidered to allow travellers to return in greater numbers, as the existing hotel quarantine system was subject to capacity constraints. Options for increasing international arrivals included reducing the length of stay in managed facilities, and/or allowing residents to quarantine at home. However, such decisions about international arrival arrangements needed to consider impact in Australia-specific settings, with diverse rates of pre-existing immunity due to vaccination and infection, different levels of community transmission and experience with stringent PHSMs. Extensive work had addressed earlier policy questions, regarding: the impact of uncontrolled outbreaks to support initial border closures in 2020 [11]; the likelihood of breaches through hotel-based quarantine pathways in the absence of vaccination [7], and; analyses of vaccination strategy and coverage thresholds that include dynamic vaccine rollout and age-specific eligibility, considering infections and hospitalisations, to support decisions for safe-reopening in the absence of arrivals [10]. The results in this manuscript were co-developed and presented to government officials as one of a suite of modelling projects helping to support data-informed decision-making. The objective of this work was to demonstrate whether and how imported cases under a variety of entry pathway assumptions would materially change the course of the local epidemiology, under differing local transmission and PHSM assumptions.

This paper reports on the linking, and use of, a series of models to estimate the consequences of infection incursions under a range of arrivals assumptions, in the context of local vaccine coverage, public health measures, and epidemiology. The goal of this analysis was to consider vaccination coverage targets and the levels of public health and social measures required to mitigate transmission in the presence of different arrival pathways and increased travel volumes following reopening. Findings from this analysis, in conjunction with work described above, informed Australia’s national reopening strategy in late-2021.

## Methods

We used a series of models to 1) estimate the likelihood that infected travellers arriving into a quarantine system would either fail to be detected or transmit infection to a worker or another traveller throughout their arrival pathway, and 2) the impact of arrival pathways on subsequently seeding infection in the community, with variable consequences depending on local epidemiology, vaccine coverage and population behaviour.

There are three distinct components of our model framework, visualised in Figure 1:

1. The *quarantine model* is an individual-based model that simulates progression through alternate quarantine pathways with differing detection effectiveness. This model quantifies the frequency and infectiousness of individuals entering the community per unit time for a given arrivals volume. A linelist of breach events from this system is generated, including information on infectiousness and residual infectious duration of each breach, based on their time since infection, and individual-level information such as vaccination status and age.
2. The *linking model* takes the breach linelist from the quarantine model (generated for a fixed source prevalence and number of arrivals), and samples the quarantine model output to generate imputed linelists of infectious individuals from the quarantine system who will enter the community over time. Existing simulations in the breach linelist are used to generate the imputed linelist for varying source prevalence and numbers of arrivals, and can combine multiple quarantine pathways.
3. The *community model* is an individual-based model that simulates transmission in the general community resulting from the introduction of infectious individuals from the quarantine system. It incorporates the impact of public health and social measures, test-trace-isolate-quarantine system effectiveness, and age-specific vaccination coverage.

**Fig 1.**
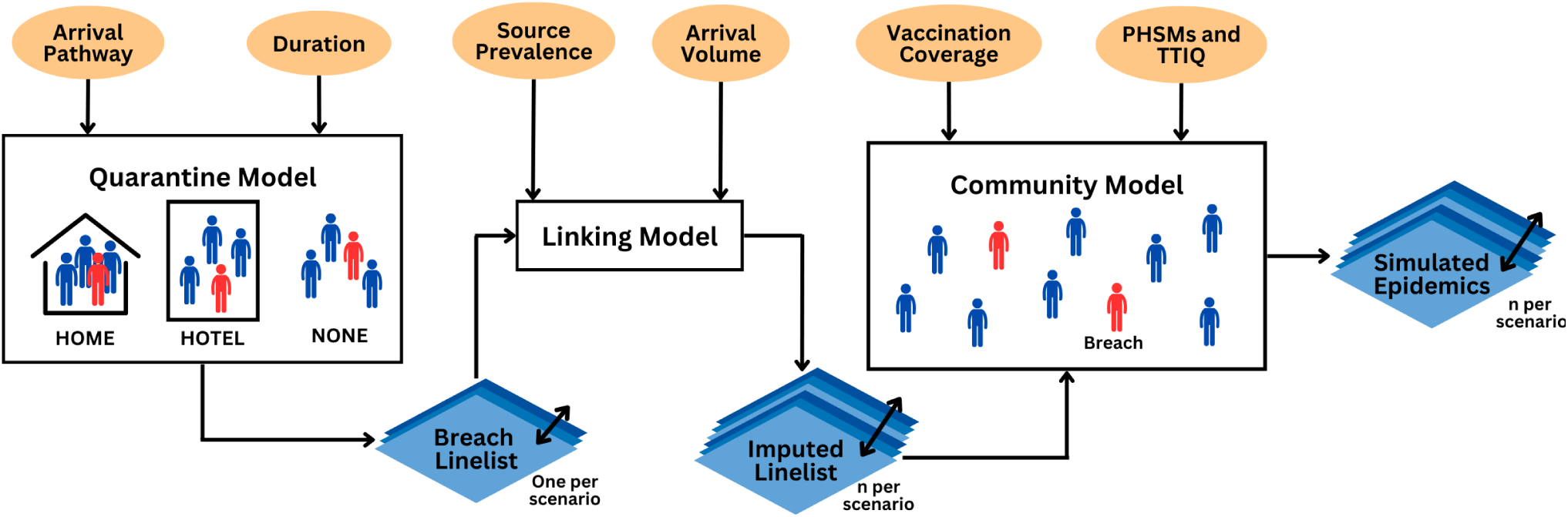
Schematic depicting the modelling framework used in this analysis. An individual-based model of the quarantine system, exploring quarantine pathway, duration and testing, is implemented to generate characteristics of breach events resulting from travellers and workers. A linelist of breach events, containing individual-level characteristics, for each scenario is generated from the quarantine model. These breach events are subsequently sampled appropriately given travel volumes and source prevalence to generate multiple linelists of breach events for each scenario, including the time the breach enters the community. Community characteristics, such as level of public health and social measures (PHSMs), test-trace-isolate-quarantine (TTIQ) system effectiveness, and age-specific vaccination coverage are implemented in the community model where breaches are introduced and subsequent outbreaks can be simulated for each breach linelist.

The quarantine [7] and transmission [10] models have been described previously, though we provide brief descriptions of the key components below and extensions relevant to this work. Code to reproduce all analyses is available at: https://doi.org/10.5281/zenodo.17719102.

### Quarantine model

The effect of quarantine systems on the number of infectious individuals entering the community from overseas, and the time during their infection at which they are released into the community, is simulated explicitly through the use of an individual-based model. The model represents individual-level time-varying infectiousness and test-sensitivity (see Supplementary Material S1.1), and models the management of individuals in quarantine and isolation including regular testing. The core features of this model have been described in detail in previous work [7], and in the following we describe the extensions for this study.

### Traveller groups

As in [7], we assume that all travellers arrive in groups of size four, and that there is a risk of transmission within the travelling party. Transmission can also occur to quarantine workers or other traveller groups within the same facility, though at a lower rate than within groups. Individuals who are identified as infected while in quarantine are removed and isolated for 10 days in separate facilities staffed by medically qualified personnel. Their accompanying travellers’ quarantine duration is ‘reset’ for another 14 days from the point of detection. The number of workers in the system scales with the numbers of travellers according to a fixed ratio (here, assumed one worker per five travellers).

A key point of differentiation compared to previous work [7] is the inclusion of family groups in which two of the individuals are children. We assume that family groups make up 18% of all arrivals, based on estimates provided by government colleagues. For such groups, the isolation of confirmed infections takes into account the constraint that children must remain in the company of at least one parent at all times. Various isolation and quarantine scenarios based on different family infection patterns are described in detail in the Supplementary Material S1.2. Arrival into Australia at the time was contingent on adults being fully vaccinated (i.e., received all required doses of a recognised vaccine product), though this was not the case for children under 12 years, for whom no vaccine was approved in Australia. As such, we assumed that all adults in the system were vaccinated, and the children (who only appear in family units) were unvaccinated. Though children were unvaccinated, they were considered to be intrinsically less infectious than unvaccinated adults (assumed 40% less infectious [12, 13]).

### Quarantine environment and duration

We consider three types of quarantine arrangements: (1) quarantine in a hotel, as exemplar of a managed facility (2) quarantine at home, and (3) no quarantine. In hotel quarantine, compliance is assured (i.e., 100%), but transmission is possible to all travellers and workers present in the facility. In home quarantine, transmission is possible only within the small group who quarantine together, but compliance is imperfect, and each day there is a probability of interaction with the general public (compliance in home quarantine is assumed 90% throughout, i.e., each individual in home quarantine had a 10% chance of interacting with the general public on each day they are in quarantine). The effectiveness of a 14-day benchmark quarantine duration is compared with only 7 days for each of these settings. The “no quarantine” scenario is implemented in the same way as home quarantine, but with a compliance probability of zero to establish a baseline absolute risk of importation without any controls measures in place, except testing of arrivals.

### Testing strategies

In all quarantine scenarios, individuals are tested via PCR upon symptom onset. If positive, cases are isolated and contacts of cases undergo an extended quarantine period within a facility (e.g., a “medi-hotel” or pseudo-clinical environment staffed by medically-trained personnel). Upon arrival into quarantine, travellers are tested on days 1, 5 and 13 for 14-day quarantine, or on days 1 and 5 for 7-day quarantine. Individuals arriving without quarantine requirements are tested on days 1 and 5. In all cases, we assume a fixed one-day delay between test and the result being known (and thus direction to, e.g., isolate if positive).

### Bridging between the Quarantine model and Community transmission model

The quarantine model outputs a time series of infected travellers and workers who have ‘breached’ the system for a fixed number of arrivals. Information on breach events includes time spent outside quarantine, relative infectiousness depending on time since infection, symptom and vaccination status, and age category (adult or child).

In order to use these linelists as input to the community transmission model, we transform and sample from them to generate an *imputed linelist*, which scales the distribution of time between subsequent breach events according to relative travel volumes and prevalence of disease in the source location. That is, given a target number of arrivals, *a_t_*, across the same timeframe as simulated from the quarantine model and a baseline prevalence of disease in the source country, *p_s_*, calculate the scaling factor,

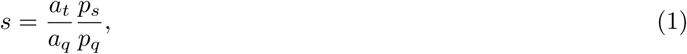

where *a_q_* and *p_q_* are the number of arrivals and the assumed prevalence from the quarantine model simulations.

Then, we calculate the time between breach events in the imputed linelist by scaling the time between breach events by *s*. To do this, we take the vector of times between breach events from the quarantine model and divide by the scaling factor *s*, effectively assuming that the quarantine model is a stationary process (e.g., [14]). This is appropriate for all quarantine systems, as the number of workers in a hotel quarantine system are assumed to be proportional to the number of arrivals into the system.

The number of breaches and their (appropriately scaled) times are then sampled over the simulation horizon. In the context of hotel quarantine, both traveller and worker breach events are sampled and combined into a single linelist of breach events. The imputed linelists of breach events are incorporated into the individual-based model as ‘pseudo’ individuals. These individuals are not part of the modelled vaccination rollout (which defines the transmission model population), but are introduced into the population at the time of breach with the properties defined by the quarantine model. As the number of breaches (hundreds) is relatively small compared to the size of the community into which they arrive (millions), these extra individuals have very little impact on population-dependent quantities such as the probability of contact.

The advantage of this imputation procedure over re-running the quarantine model is the time to produce results. The quarantine model captures considerable detail on a relatively small system, but as a result, the computation time becomes large for a given number of quarantine arrivals. The imputation procedure efficiently generates a series of linelists accounting for different travel volumes and/or prevalence in the source population. This allows for greater exploration of the number of arrivals and, while we only considered each pathway in isolation in this work, different combinations of arrival pathways can be readily configured.

### Community transmission model

A separate individual-based model is used to model transmission of SARS-CoV-2 in the general community. This model has been described in detail previously [10], and the key features are summarised here.

The transmission pathway in this model is Susceptible-Exposed-Infectious-Recovered (SEIR), modified by vaccination. Individuals in the exposed class (E) are infectious but are not currently displaying symptoms. After their exposed period, individuals are classified as either symptomatic or asymptomatic. Individuals who are recovered are considered permanently immune given the timescale of several months relevant to the reopening agenda, although evidence of waning infection protection was emerging in late 2021 [15], with additional evidence generated since (e.g., [16]). The number of contacts made by each infectious individual is Negative Binomial distributed (with the exception of those in isolation, who have zero contacts by definition). That is, the number of contacts an individual in age group *i* makes with individuals in age group *k* is,

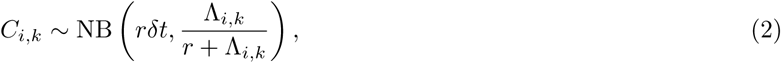

where *r* is the dispersion parameter, *δt* is the size of the current time step and Λ*_i,k_* is the (*i, k*)^th^ element of the contact matrix, **Λ**. The *C_i,k_*contacts are chosen uniformly from individuals of age *k* in the population (some of whom may not be susceptible). The probability of infection given contact with a susceptible individual is,

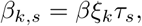

where *β* is the baseline probability of infection given contact, *τ_s_* is the relative transmissibility of the infector (*s* = 1 for symptomatic, *s* = 0 for asymptomatic), and *ξ_k_*is the relative susceptibility of the infectee (which depends on their age group). The probability of an infected individual having symptomatic disease is age-dependent, as described in [10] based on estimates from [12]. Younger individuals have the lowest probability of symptomatic disease, which increases with age.

Vaccination is incorporated into the transmissibility of the infector, such that,

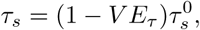

with 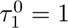 for symptomatic, and 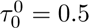 for asymptomatic individuals and *V E_τ_* is the vaccine efficacy against onward transmission. Vaccination of the infectee also modifies their relative susceptibility, such that,

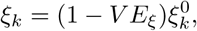

where 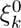 is the baseline susceptibility for an individual in age bracket *k*, and *V E_ξ_*is the vaccine efficacy against infection.

### Vaccination coverage

The proportion of each age group that received a vaccine is taken from modelled estimates of the vaccination rollout used as part of Australia’s national reopening plan for COVID-19 [10]. Briefly, an agent-based model informed the anticipated vaccine rollout, incorporating information on location and allocation data on vaccination sites and location data for the Australian population. A fraction of the population in each geographic location that satisfied the eligibility criteria over time would seek a vaccine. Sites were allocated a stock of vaccine, and would administer vaccines within that capacity to those seeking vaccination. The majority of vaccines deployed in Australia were mRNA vaccines, following early use of the ChAdOx1 vaccine in older age groups. For the purposes of this work, vaccination coverage inputs were taken as a fixed snapshot of this rollout at 70%, 80%, or 90% of the eligible population having received two doses, and a small percentage thus having only received one dose. The vaccination coverage within each age group across each vaccine product is taken from the observed and modelled rollout up to each vaccination threshold. This choice was made to simplify the interpretation, and attribute differences to the differential public health and social measures or arrival pathways. The one- and two-dose coverage by age group and vaccine product corresponding to the different population coverage levels is shown in Supplementary Figure S1.

### Effect of public health and social measures and test-trace-isolate-quarantine

The effects of public health and social measures (PHSMs) and the test-trace-isolate-quarantine (TTIQ) system are captured directly within the community transmission model. The impact of PHSMs is captured as a reduction in *R*_0_ (or equivalently transmission potential, as described in Ryan *et al.* [17]), in line with previous work [10]. We consider two sets of PHSMs: ‘baseline’, representing population distancing behaviour under minimal density/capacity restrictions and no major outbreaks, and ‘low’, representing additional capacity limits on workplaces, retail and recreational activities. The impact of these restrictions on transmission was calibrated based on behavioural and case data from 2020 and 2021 in New South Wales, Australia [18].

The effect of TTIQ is incorporated by modifying the individual’s probability of transmission. In particular, an infected individual’s probability of transmission is set to zero when they are directed to isolate (i.e., assuming 100% compliance). The time at which this occurs is taken from a distribution of times from infection to isolation, described in full in [18]. We use two different distributions, termed ‘optimal’ and ‘partial’ TTIQ. These distributions were estimated from two distinct epidemiological contexts in two Australian jurisdictions during 2020–2021. The optimal delays were estimated where caseloads were relatively low (tens of cases per day) and TTIQ was clearly suppressing transmission, whereas the partial distribution was estimated where caseloads were considerably higher (hundreds of cases per day) and contact tracing systems were reported as under stress [18]. The median delays from infection to isolation are approximately 3.44 days (optimal) and 8.95 days (partial) — full distributions are shown in Supplementary Figure S2.

### Simulated scenarios

#### Arrival setting

In the following, we consider two different settings in which the arrivals are introduced — one where there is no existing transmission, and one where there are relatively low levels of existing transmission — termed ‘no existing transmission’ and ‘existing transmission’ herein. The level of PHSMs and TTIQ are assumed to differ in these settings. Australian jurisdictions that had experienced substantial outbreaks at the time this work was completed were anticipating some level of PHSMs to persist during the reopening phase. In contrast, jurisdictions that had largely avoided substantial outbreaks were less inclined to impose these measures during the reopening phase, particularly as they had previously not experienced such measures for any significant period of time. Further, we expected that settings with no existing transmission (i.e., starting with no cases to manage) were more likely to maintain ‘optimal’ TTIQ [19], whereby the distribution of time from infection to isolation is relatively short. Where there is existing transmission, we assume that only ‘partial’ TTIQ is sustainable given the case loads requiring management. While the ability to maintain these TTIQ efficiencies is dependent on the number of cases a public health unit is managing at a given time, we did not incorporate this dynamic feedback in the model. Instead, we assumed that the TTIQ efficiences were maintained throughout, and describe the expected differences where they would not be expected to be sustained (i.e., when daily new infections were high) or where improved performance is likely (i.e., daily new infections were low).

#### Quarantine pathways

We compare the 7- and 14-day home and hotel quarantine strategies against two reference scenarios — no quarantine (but testing on days 1 and 5) and a 14-day hotel quarantine scenario where all individuals are unvaccinated, representing the state of the system prior to the national reopening plan. We collectively refer to the quarantine type and duration as the ‘arrival pathway’ herein.

#### Source prevalence

Given variable infection prevalence and exposure risk across source countries, and mandated pre-embarkation testing requirements, we assumed a fixed 1% likelihood that any unvaccinated traveller would arrive infected, having not been detected in the origin country pre-departure or having acquired infection in transit. Vaccination is assumed to provide 80% protection against infection given exposure, reducing the proportion of infected vaccinated arrivals to 0.2%. These assumptions on source prevalence are fixed for all scenarios.

#### Community vaccination coverage

In each case, vaccination coverage in the community is fixed at 80%. To explore the impact of vaccination coverage on transmission in each setting, we consider breaches from one arrival pathway (7-day home quarantine with 90% compliance) where community vaccination coverage is 70%, 80% and 90%, on the basis that the relative ordering of the arrival pathways will be consistent.

#### Arrival volume

To consider the implications of increasing arrivals, future projections were benchmarked against pre-pandemic (i.e., 2019) travel volumes of Australian citizens and permanent residents, who were the highest priority returns, including the proportion of children. The number of arrivals into the communities is informed by arrival volumes into the Australian states of New South Wales and Western Australia, as representative large- and moderate-sized jurisdictions, with their corresponding population size (approximately 8 and 2.6 million, respectively). As a result of these demographic differences, the magnitude of results should not be compared across these settings. The number of arrivals into the communities is fixed at 40% of the average weekly pre-pandemic arrivals (i.e., using total arrivals from 2019). This corresponds to approximately 10,000 weekly arrivals in the setting with no existing transmission, and 33,000 weekly arrivals in the setting with existing transmission. Arrivals are initiated at day 40 in scenarios where there is existing transmission, and day 0 otherwise. The impact of increasing arrival volumes from 40% to 80% of pre-pandemic baseline is explored in the context of one arrival pathway, shown in Supplementary Material S2.

#### Simulations

For each scenario, we generate 200 simulations, each with a time horizon of 500 days. Results from the community transmission model are presented in Figures 3–7 as 50% and 90% intervals of daily infections over the time horizon. Supplementary results S2 show figures and tables of the infections relative to a baseline for each scenario.

**Fig 2.**
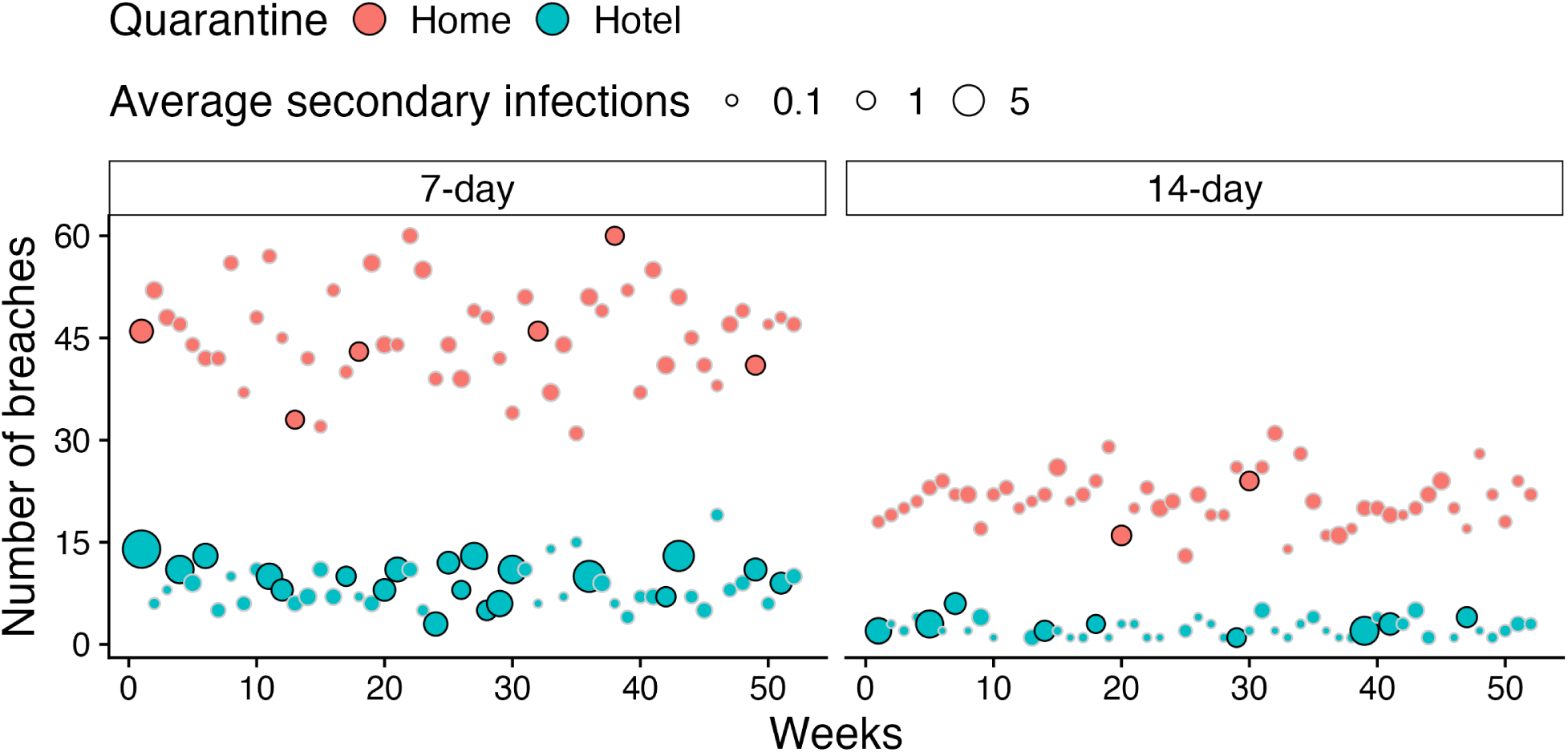
Number of breaches from the quarantine system and the average number of secondary infections caused by the breaches by week for one year, for one simulation from the 7- and 14-day duration for home and hotel quarantine. Weeks where the average secondary infections across all breaches is greater than 1 have a solid black outline. These results assume 40% pre-COVID arrival volume into a larger jurisdiction (New South Wales).

**Fig 3.**
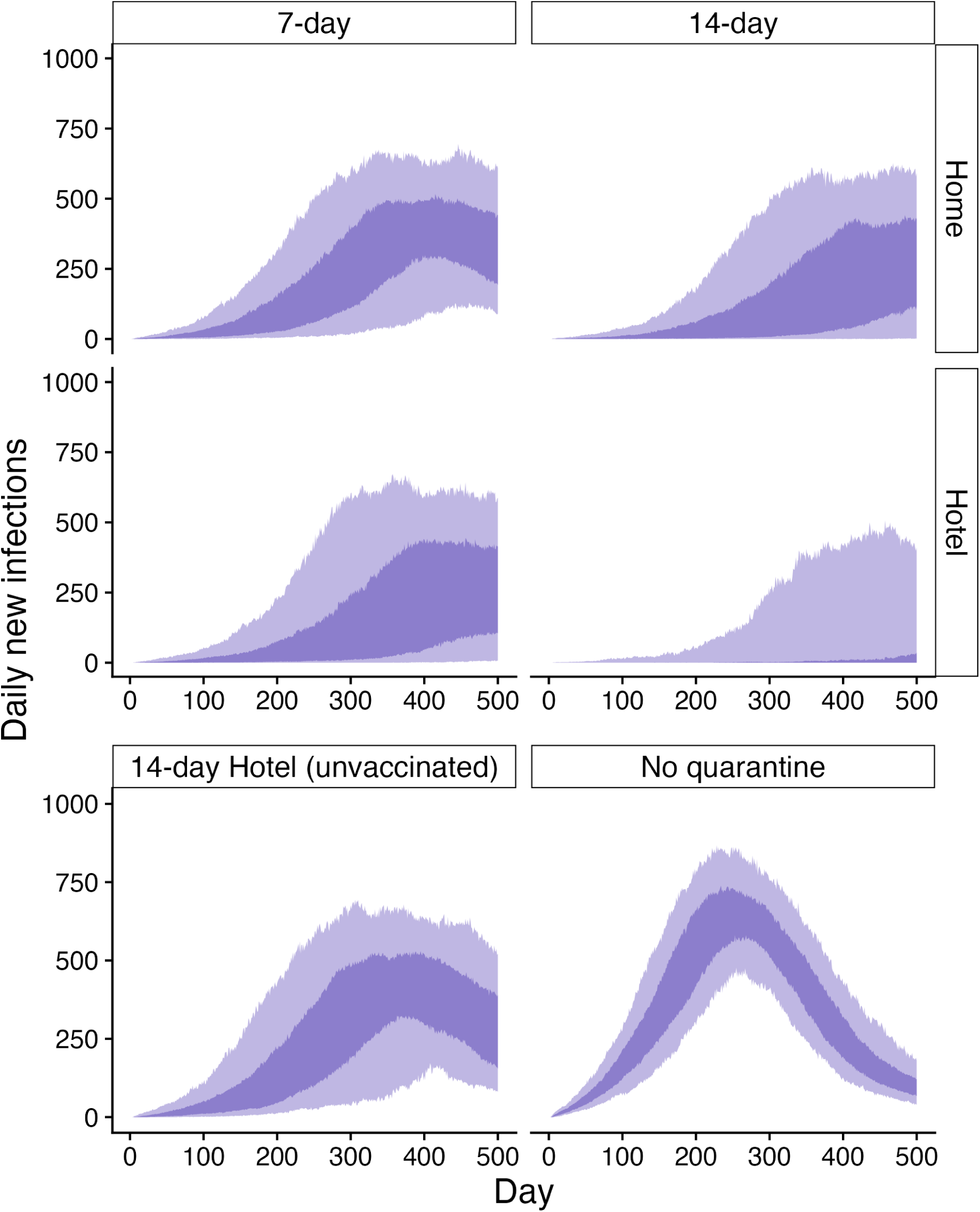
Daily new infections in a community with no existing transmission as a result of breaches via 7- or 14-day home or hotel quarantine (top panel). No quarantine or 14-day hotel quarantine (all arrivals unvaccinated) are shown in the bottom panel. Baseline PHSMs and optimal TTIQ are in place in the community, and vaccination coverage is fixed at 80%. All infections in the community are seeded by breaches from the arrival pathways. Dark and light ribbons represent 50% and 90% intervals, respectively.

**Fig 4.**
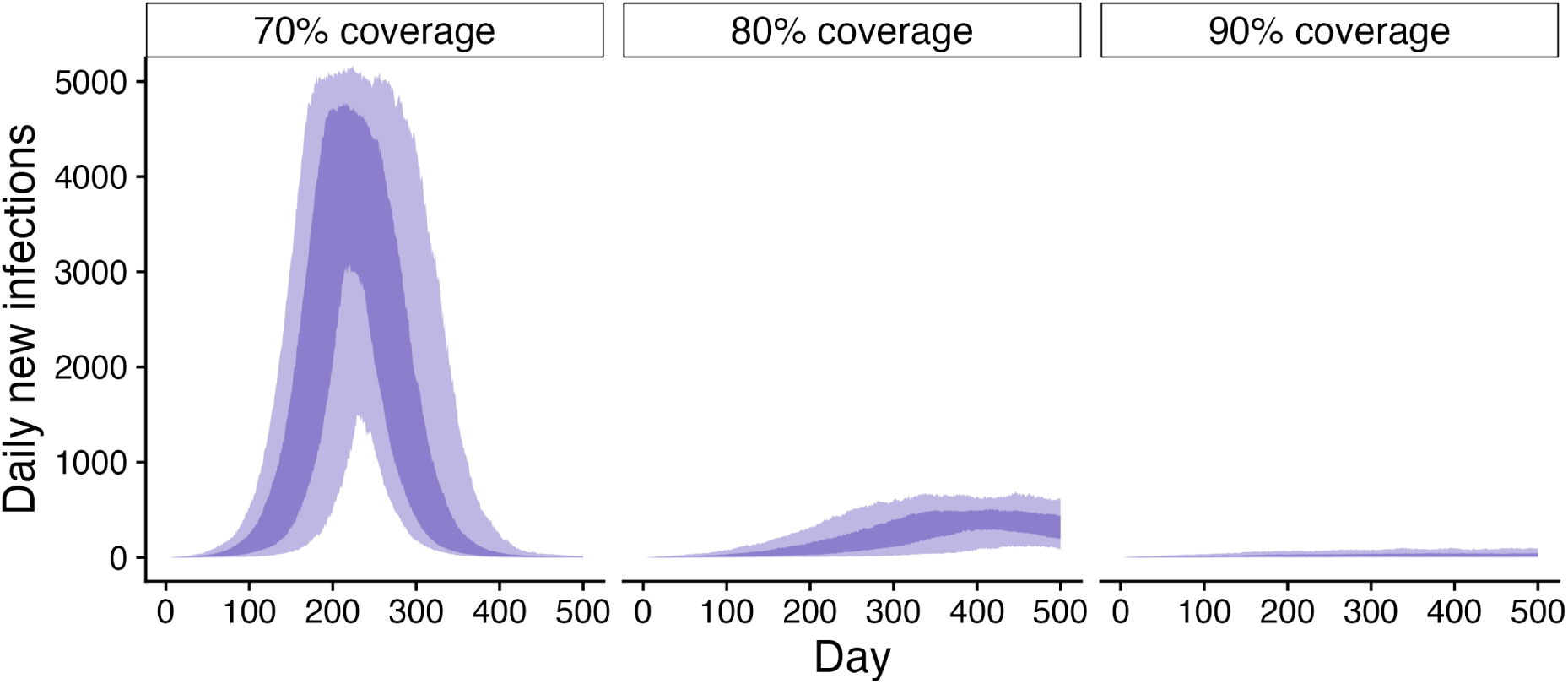
Daily new infections as a result of imported infections via 7-day home quarantine into a community with no existing transmission with low PHSMs and optimal TTIQ, where community vaccination coverage is 70% (left), 80% (middle) or 90% (right). Arrival volumes are 40% of 2019 levels. All infections in the community are seeded by breaches from the arrival pathways. Dark and light ribbons represent 50% and 90% intervals, respectively. Note that the 80% coverage results shown here are the same as those shown above in Figure 3.

**Fig 5.**
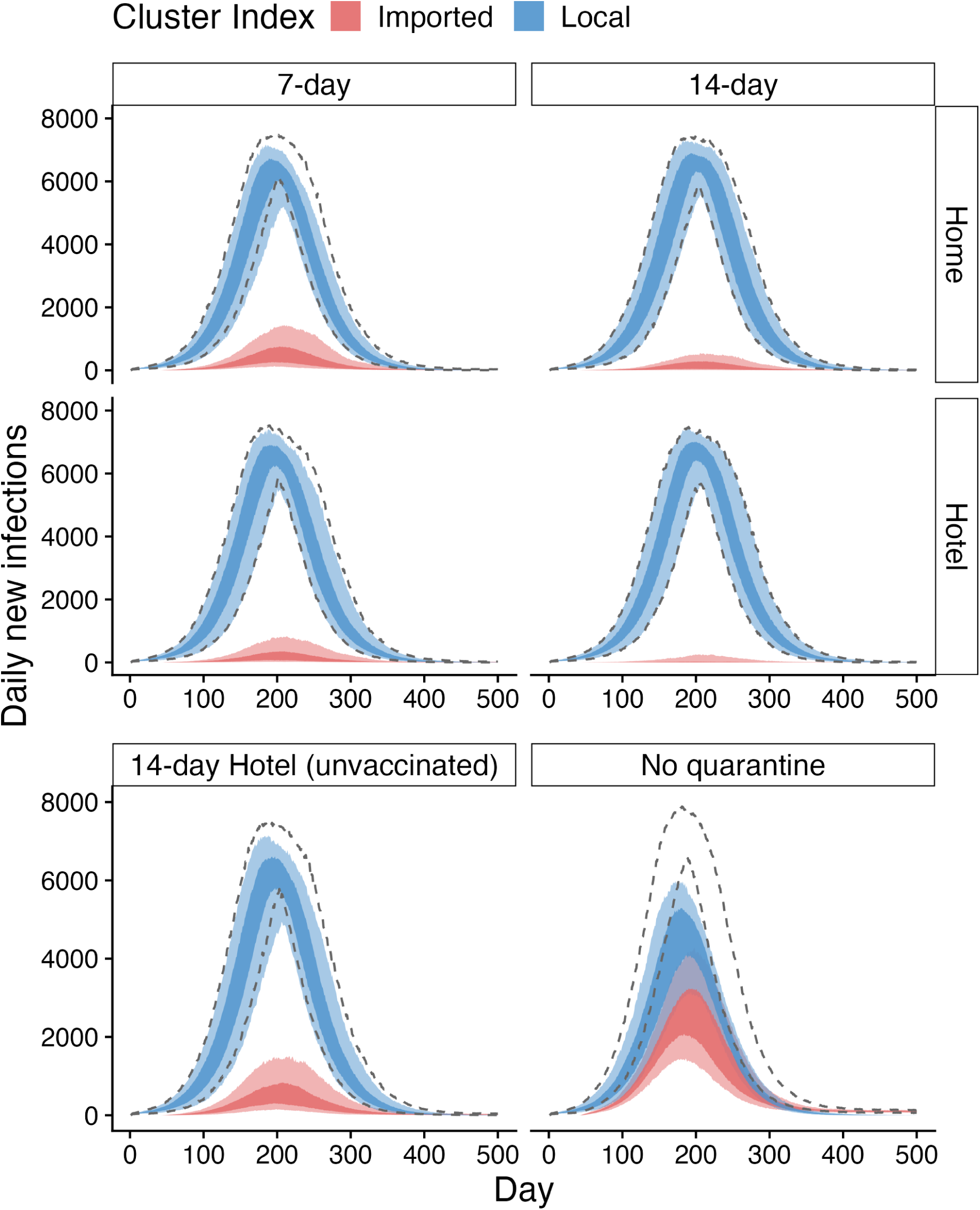
Daily new infections in a community with existing transmission as a result of infections imported via 7- or 14-day home or hotel quarantine (top panel). No quarantine and 14-day hotel quarantine for unvaccinated arrivals are shown in the bottom panel. Baseline PHSMs and partial TTIQ are in place in the community, and vaccination coverage is fixed at 80%. Colour represents whether the outbreak was seeded by a locally derived infection, or an imported infection. Dark and light ribbons represent 50% and 90% intervals, respectively. The dashed black lines represent the 90% intervals for the total infections (i.e., the sum of infections seeded by local and imported infections). Arrivals initiate on day 40.

**Fig 6.**
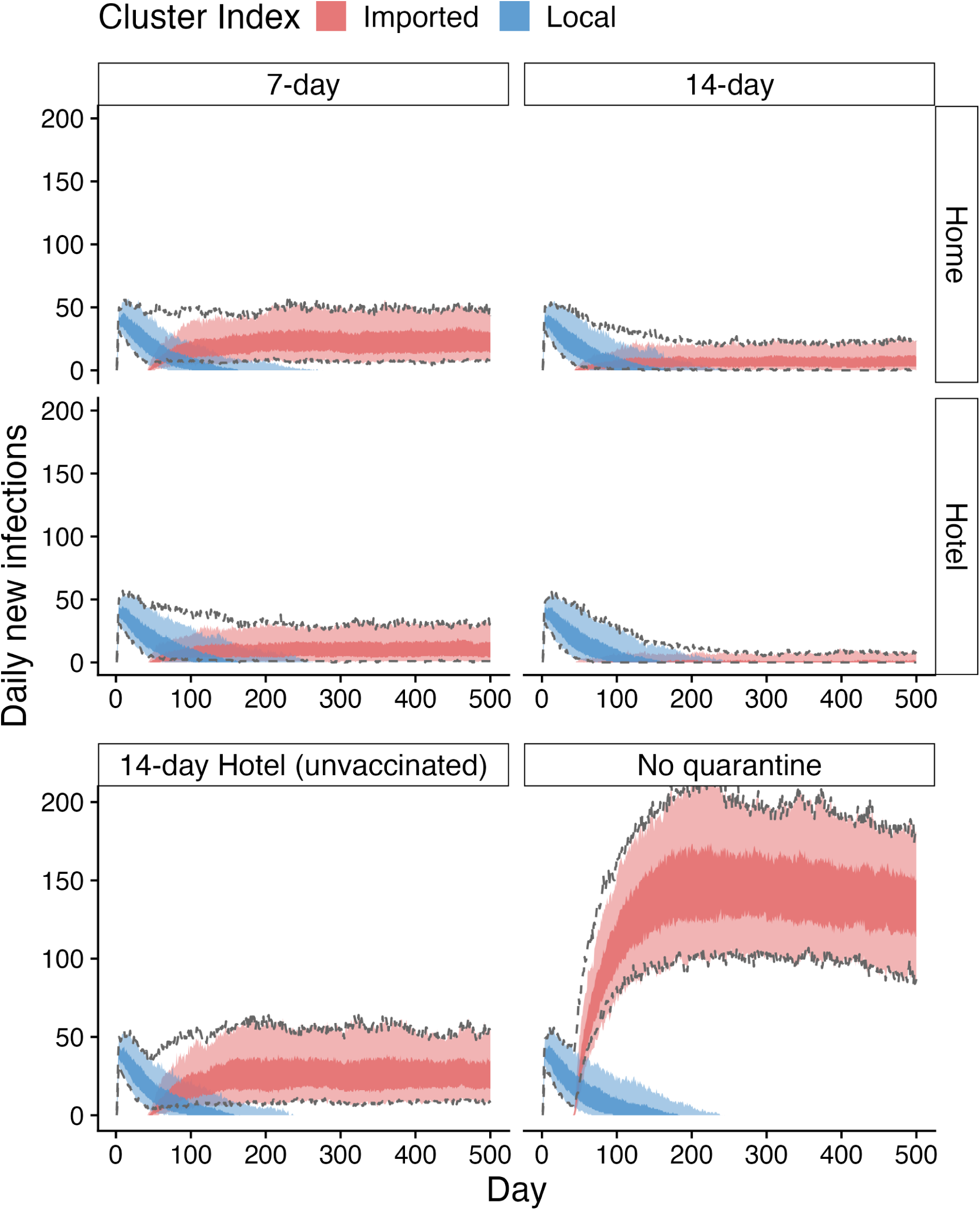
Daily new infections in a community with existing transmission as a result of infections imported via 7- or 14-day home or hotel quarantine (top panel). No quarantine and 14-day hotel quarantine for unvaccinated arrivals are shown in the bottom panel. Low PHSMs and partial TTIQ are in place in the community, and vaccination coverage is fixed at 80%. Colour represents whether the outbreak was seeded by a locally derived infection, or an imported infection. Dark and light ribbons represent 50% and 90% intervals, respectively. The dashed black lines represent the 90% intervals for the total infections (i.e., the sum of infections seeded by local and imported infections). Arrivals initiate on day 40.

**Fig 7.**
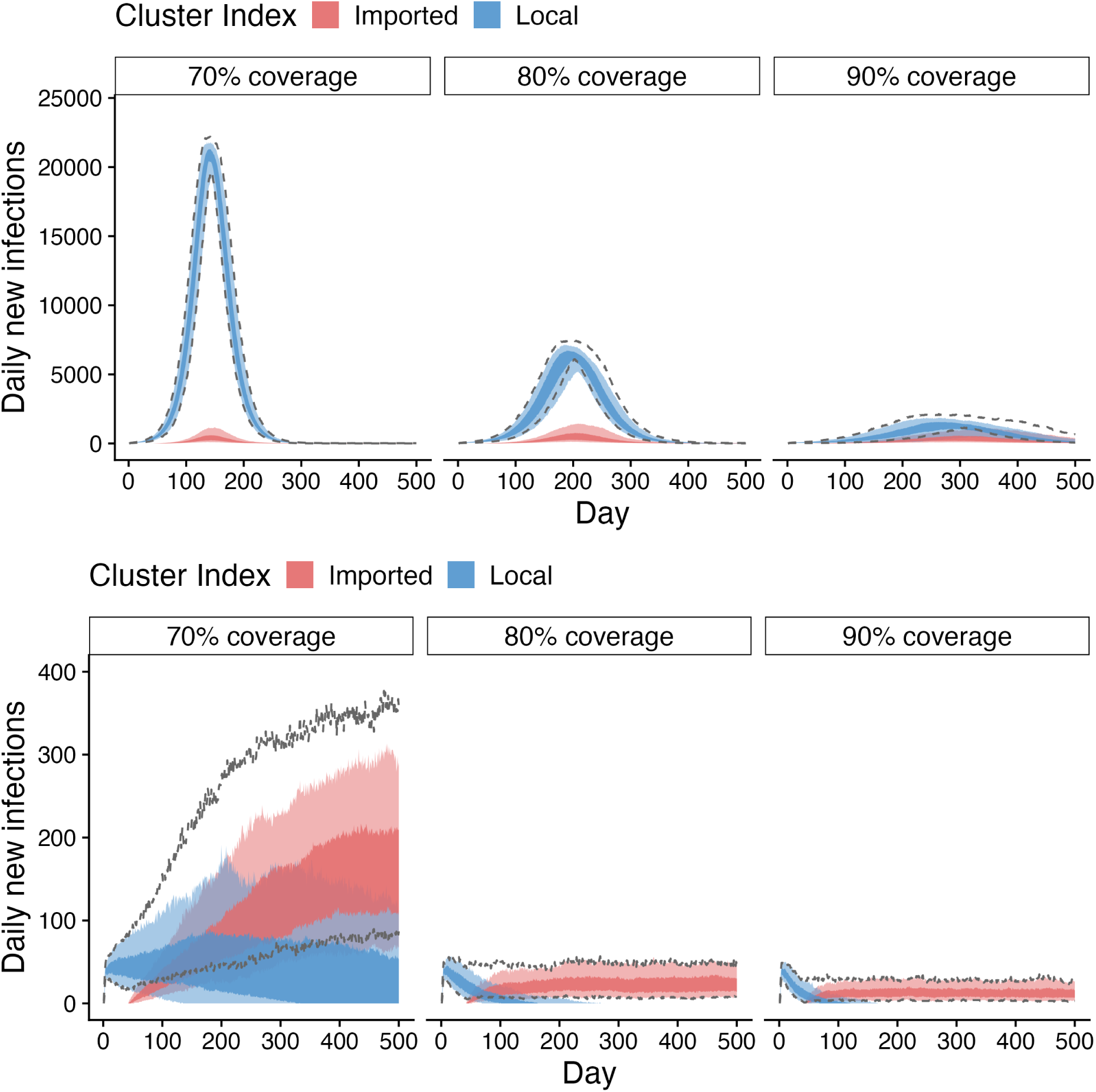
Daily new infections as a result of infections imported via 7-day home quarantine into a community with existing transmission under baseline (top) or low (bottom) PHSMs, where community vaccination coverage is 70% (left), 80% (middle) or 90% (right). Arrival volumes are 40% of 2019 levels, and partial TTIQ is in place throughout. Colour represents whether outbreak was seeded by a local or imported infection. Dark and light ribbons represent 50% and 90% intervals, respectively. The dashed black lines represent the 90% intervals for the total infections (i.e., the sum of infections seeded by local and imported infections). Arrivals to the quarantine system initiate on day 40. Note that the results for 80% vaccination coverage shown here are the same as those in Figures 5 and 6 above.

## Results

### Relative efficacy of different arrival pathways

The average characteristics of breach events per week for 7- and 14-day home and hotel quarantine with a fixed number of arrivals are presented in Figure 2. The home quarantine pathway produces consistently more breaches than hotel quarantine. However, breaches from hotel quarantine correspond to a greater average number of secondary infections (larger points), with the average infectiousness of individuals being greater than 1 more frequently (black outline). The increased infectiousness of individuals from hotel quarantine is a result of: the potential for transmission from travellers to workers, who may then spend time in the community early in their infectious period before being detected, and; transmission between traveller groups within the hotel system, with the risk of undetected infected individuals being subsequently released from hotel quarantine relatively early in their infectious period.

Whether the increased number of breaches from the home quarantine system, or the heightened infectiousness (average secondary infections) of breaches from the hotel quarantine system has a greater impact on community level infection dynamics is explored via the community transmission model.

### Transmission in context of existing epidemiology

The impact of these breaches on the overall infection dynamics is highly dependent on the state of the system at the time of the breach. For example, if transmission is well established, then the introduction of a few infectious individuals in the community may have little impact on the system dynamics. However, if there is little or no ongoing transmission in the community, then even a few breaches may have a substantial impact. Further, the level of vaccination coverage in the community, stringency of public health and social measures (PHSMs), efficiency of the test-trace-isolate-quarantine (TTIQ) system, and the characteristics of the breach events resulting from the different arrival pathways will impact the community-level infection dynamics. Herein, we present the results for each scenario, considering these differences. We include a brief description of the key results in Table 1 for easy reference.

**Table 1.** Summary of key results.

| Setting | TTIQ | PHSM | Key results |
| --- | --- | --- | --- |
| No existing transmission | optimal | baseline | Figure 3. Slow transmission over a long time-scale (i.e., reproduction number close to 1). Most epidemics are not resolved by day 500 for all arrival pathways. Low numbers and slow spread would likely be manageable by the public health response, or, would allow sufficient time to enact a stronger response to further mitigate transmission. Otherwise, the scale of infection near the peak may reduce the effect of ongoing TTIQ below the assumed level, increasing the amount of transmission that would eventuate.<br>Figure 4. Arrivals to the population with only 70% vaccination coverage is likely not sufficient to avoid a substantial outbreak. |
| Existing transmission | partial | baseline | Figure 5. Daily infections peak around 6000. This scale would likely reduce the effectiveness of TTIQ below the assumed level, further increasing the amount of transmission. Local transmission dominates any effect at reducing overall transmission by placing arrivals in the different quarantine pathways.<br>Figure 7 (top panel). Significant outbreaks are likely to eventuate should arrivals begin with only 70% vaccination coverage in the community. |
|  |  | low | Figure 6. The addition of low PHSMs reduces the reproduction number below 1, meaning transmission chains are generally short. Ongoing transmission is driven exclusively by quarantine breaches. The sustained level of transmission is directly related to the rate of breaches from the quarantine pathways, but consistently $<50$ infections per day across all arrival pathways. This low level of transmission would likely be manageable.<br>Figure 7 (bottom panel). 70% vaccination coverage leads to an increase in the daily infections up to at least 5-fold, compared to $\geq 80\%$ vaccination coverage. |
|  | Overall |  | Figures 3, 5, 6. 14-day hotel quarantine consistently results in fewer breaches and thus transmission chains introduced by arrivals, followed by 14-day home quarantine, then 7-day hotel and home quarantine. In the presence of existing transmission, minimal benefit is observed from stringent quarantine pathways. In the absence of local transmission, the local risk tolerance would dictate which arrival pathways are sufficient. The arrival volumes that home and hotel quarantine are able to manage, and the resources required to maintain each, should be factored into decisions on the best strategy. |

### Arrivals into a setting with no existing transmission

Figure 3 shows the daily number of new infections in a community with no existing transmission for each arrivals pathway. It is assumed that ‘low’ PHSMs were in place in the community and, given the absence of ongoing transmission, that optimal TTIQ is maintained (i.e., the shortest average delay between infection and notification). The results show that both forms of quarantine result in fewer infections than no quarantine in this context, across the same time frame. The least stringent quarantine, 7-day home, performs comparably, albeit marginally worse, than the 14-day hotel quarantine of unvaccinated arrivals (mean infections over time-horizon relative to ‘no quarantine’: 54%, with 50% interval [30%,76%], vs 39 [8%,67%]; Supplementary Table S3), where the 14-day hotel quarantine is indicative of the quarantine system prior to widespread vaccination. For all quarantine pathways, there are frequent incursions into the community which would require a sufficient public health response to re-establish elimination, or reduce infections to a level manageable by the jurisdictions public health units and healthcare system. However, the time frame of these simulations (500 days) is sufficiently long, and growth is sufficiently slow, that if such a scenario were to occur in reality, ongoing situational assessment would enable such a public health response.

Figure 4 shows the impact of reintroducing arrivals via the 7-day home quarantine pathway (90% compliance) where vaccination coverage in the community is 70%, 80% or 90%, in the presence of low PHSMs. These show that a substantial outbreak would likely occur in such a population should individuals start entering the community when vaccination coverage is only 70%, peaking at around 4700–5500 daily infections (approximate 5*^th^*, 95*^th^* percentiles of peak infections across simulations). Further, it is unlikely that optimal TTIQ would be maintained with infections at this level, and thus daily infections would likely peak higher, and with longer epidemics, than those shown here. Delaying the reintroduction of arrivals until 80% or 90% vaccination coverage were achieved results in drastically smaller, more manageable, outbreaks (20% [15%, 25%] and 2% [1%, 3%] for 80% and 90% vaccination coverage relative to 70%; Supplementary Table S4).

The impact of increasing arrival volumes from 40% of pre-pandemic baseline to 80% via 7-day home quarantine (90% compliance) in the presence of low PHSMs is shown in Supplementary Figure S14 (top). As expected, increasing arrival volumes marginally increases the daily number of infections by increasing the number of seeding events, but has less impact than the quarantine pathways, or the vaccination coverages considered.

### Arrivals into a setting with existing transmission

Figures 5 and 6 show the daily number of new infections in a community with existing transmission where baseline or low PHSMs are in place (respectively) for each arrivals pathway. In each scenario, partial TTIQ is assumed to be maintained throughout the duration of the simulations. Given the two sources of infection (arrivals and local) these figures distinguish the daily new infections by the source of the transmission chain (red and blue, respectively), and show the 90% intervals for the total daily infections (black dashed lines).

In the presence of baseline PHSMs (Figure 5), there are negligible differences in the total infections across quarantine pathway and duration. The largest reduction in overall infections compared to no quarantine is 2.4% [1.4%, 2.8%] for 14-day hotel quarantine (Supplementary Figure S10 and Table S5). This is due to the fact that the number of incursions (and thus new chains of transmission) is very low relative to the established community transmission. The difference between the source of daily infections (arrival or local) is smaller in the absence of quarantine.

With low PHSMs, local transmission can effectively be eliminated (Figure 6). The only driver of infection is incursions from arrivals — the low PHSMs are sufficient to mitigate the relatively low number of secondary infections from each infectious arrival — resulting in a stable number of daily infections over the time horizon. The relative number of infections across the time-horizon is directly proportional to the breach characteristics of each quarantine pathway (Supplementary Figure S11 and Table S6). It should be noted that this constant level of daily new infections is not from a single incursion that continues over time. Rather, it is frequent, short chains of transmission that become extinct, only to be replaced by new chains of transmission seeded by infected arrivals. The low burden on the public health system would likely correspond to improved TTIQ performance than the assumed partial TTIQ distribution.

The relatively small contribution of arrivals is consistent across population vaccination coverage. Figure 7 shows how epidemic dynamics in the community with either baseline (top) or low (bottom) PHSMs would change were arrivals introduced (via 7-day home quarantine with 90% compliance) at 70%, 80% or 90% vaccination coverage. As expected, higher vaccination coverage results in a substantial decrease in the number of daily infections, though the pattern of sustained transmission vs frequent imports remains the same. In particular, with baseline PHSMs in place, the number of infections seeded by arrivals remains relatively low as local transmission dominates infection dynamics. With low PHSMs in place, vaccination regulates the magnitude of the equilibrium level of infections reached over this time horizon (Supplementary Figures S12 and S13 and Table S7).

Supplementary Figure S14 shows the impact of increasing arrival volumes from 40% of pre-pandemic baseline to 80% in the community with baseline (middle) or low (bottom) PHSMs, via the 7-day home quarantine (90% compliance) pathway. As expected, the arrival volume has a minimal impact on the infection dynamics — which are otherwise dominated by local transmission for baseline PHSMs, or simply moderates the equilibrium daily infections over the time horizon when low PHSMs are in place.

## Discussion

Quarantine breaches are low probability events with the potential for substantial community consequences. To explore both the frequency and scale of outbreaks arising from arriving travellers, we developed a model framework that represented both quarantine and community transmission in a consistent, unified framework. The quarantine model included a detailed representation of time-varying infectiousness and test sensitivity within individuals, enabling a precise characterisation of both the probability of a traveller leaving quarantine while still infectious, and their level of infectiousness at that time. The community transmission model reflected the heterogeneity in vaccination coverage, arrival volumes, and PHSM and TTIQ settings that were present across Australian jurisdictions.

### Results from this study

Our modelling study demonstrated the need for high vaccination coverage thresholds (i.e., 80% of the eligible population) to maintain SARS-CoV-2 infections at manageable levels following the introduction of a large volume of arriving travellers, given the epidemiology in Australia in late-2021. This result held regardless of whether there was existing transmission in the community when borders were reopened. At lower levels of vaccine coverage (e.g., 70% of the eligible population), the ongoing use of more stringent PHSMs would be required to maintain infections at manageable levels (Figure 7).

We found that the impact of quarantine varied depending on the epidemiological characteristics of the jurisdiction. In communities that were free of SARS-CoV-2 and had only baseline PHSMs in place, our analyses showed that the arrivals pathway could have a meaningful impact on infection dynamics in the community following reopening (Figure 3). As expected, a longer quarantine duration corresponds to reduced infections and a later peak, as quarantine breaches occur less frequently and involve less infectious travellers (Figure 2). In contrast, ongoing quarantine has minimal impact when there is existing community transmission and only baseline PHSMs (Figure 5). In this scenario, transmission is sustained as a consequence of existing local cases, and quarantine breaches have little impact. The total number of infections over the course of an outbreak were near-identical for each quarantine pathway, with only the source of the transmission chains differing according to the force of infection from the quarantine pathway (Supplementary Figure S10, Table S5). Similar conclusions hold for the communities with existing transmission and low PHSMs. In this case, the combination of vaccination coverage and PHSMs maintain the reproduction number below the critical threshold of 1 from early in the simulated period such that transmission is not sustained in the community from local or imported infections. The magnitude of the sustained daily infections is proportional to the force of infection arising from the quarantine pathway, where more effective strategies (e.g., 14-day hotel quarantine) result in lower sustained daily infections than less effective strategies (e.g., no quarantine). As expected, increasing arrival volumes generally leads to an increase in the number of breach events and subsequent community infections (Supplementary Figure S14), except where existing local transmission dominates infection dynamics in the presence of baseline PHSMs (middle panel). In that scenario, ongoing transmission chains have a greater impact than the scale of arrivals and breach events as the increase in infections from breach events is linear, but growth from transmission within the community is exponential.

### Australia’s national re-opening plan

In conjunction with other analyses [10, 17], this work directly informed vaccination coverage targets as part of Australia’s national reopening plan [20], and contributed key information to support national policy on international travel requirements. The national mandate on quarantine for overseas Australian residents returning to Australia was removed from 1 November 2021 [21], however, the exact timing quarantine requirements were removed differed by jurisdiction. New South Wales (NSW) was one jurisdiction in late 2021 with ongoing community transmission and stringent PHSMs in place (e.g., [22], [23]). Cases in NSW were on the decline into October, as 80% two-dose vaccination coverage was reached on 16 October 2021 [24] and the state was onboard with reinitiating international travel from 1 November [25]. In contrast, Western Australia, which had experienced relatively little local transmission since the beginning of the pandemic, favoured higher vaccination coverage – targeting 90% – prior to reopening [26]. The state initially announced in December 2021 that they would reopen borders to fully vaccinated arrivals from interstate and overseas on 5 February 2022 [27], though this was ultimately delayed in January 2022 following concerns about Omicron [28].

### Related modelling studies

Several other studies have separately explored the impact of arrivals: the risk of importation through different quarantine pathways, duration, and in the presence of different testing strategies [7, 29–31], or the risk of outbreaks per infected arrival introduced to a community [30, 32, 33]. Our findings are consistent with those reported in Leung *et al.* [34] and Vattiato *et al.* [35], who similarly explore the impact of infected arrivals introduced to a community, and the impact of PHSMs and TTIQ in the presence of varying vaccination coverage. Leung *et al.* [34] explored vaccination coverage thresholds that would be required to avoid an outbreak for a range of vaccination efficacy assumptions. Quarantine and testing of international arrivals consisted of testing on arrival with the test result indicating isolation, or quarantine ranging from 1–14 days. The risk of transmission amongst travellers and to workers, and the isolation of infected travellers detected while in quarantine, was not considered. They showed, consistent with our analyses, that imposition of some control measures would be required to supplement achievable vaccination coverage targets, and that maintaining quarantine and testing of arrivals would be necessary to reduce the risk of outbreaks until high vaccination coverage in the community was achieved. Similarly, Vattiato *et al.* [35] explored dynamic management strategies in order to maintain health system capacity within defined limits, and quantify the anticipated amount of time with different levels of restrictions similar to the static analysis conducted in Australia [17]. With a focus on the community level implications of SARS-CoV-2 transmission and restrictions required to maintain health capacity rather than specific border policies, reasonable simplifying assumptions were made regarding infected overseas arrivals — fixed numbers of infected arrivals per year and efficacy of quarantine that acted on each infected arrival independently. The results of this analysis are also consistent with those presented here, concluding that infected arrival volumes had negligible impact on health burden once a high vaccination coverage was achieved and community transmission was widespread.

### Limitations

As with any modelling study, the work here is subject to limitations based on knowledge at the time of the analysis, and necessary simplifying assumptions. The transmission rate and vaccine efficacy parameters were specific to the Delta variant, which was the most prevalent variant globally at the time of analysis in September–October 2021 — prior to the emergence of Omicron.

The emergence of the Omicron variant, with different transmission and vaccine efficacy properties, combined with a shifting political landscape, mean that the scenarios modelled here do not represent what ultimately occurred during the reopening period. As such, retrospective analyses of the modelled scenarios performance have not been considered. The immune-evading properties of Omicron would result in considerably different outcomes than those presented here. Increased transmissibility, and the modest impact of vaccination on transmission and susceptibility [8, 36], would be expected to drive larger outbreaks in all scenarios. Further analyses, beyond the scope of this work, would be required to establish under which circumstances this would increase the reproduction number above the critical threshold of 1, where transmission was otherwise not sustained for Delta. The larger and likely more rapid outbreaks would necessitate more rigorous constraints to mitigate transmission. However, given the considerably lower pathogenicity of Omicron to earlier variants, particularly in a highly vaccinated population [36], this level of restriction would have been disproportionate to the public health risk.

The risk assessment reported here was conducted in conjunction with related model-based analyses of vaccination allocation strategy and target thresholds in the Australian context [10, 17]. In contrast to [10], which estimated clinical and mortality outcomes in the context of the Delta variant for all of Australia, the present analysis focused on the relative differences in infection dynamics between vaccination thresholds and quarantine pathways. For further context regarding the anticipated clinical or mortality burden associated with the scale of infections presented here, we direct the reader to [10], or jurisdiction-specific analyses conducted at the time in [37].

As in other model-based analyses of borders and quarantine (e.g., [33]), we assumed static vaccination coverage to simplify the interpretation of observed differences in our simulation results. As explored in more detail in [10], the vaccination program continued to rollout rapidly across Australia during late 2021 and into 2022, but with substantial variability across jurisdictions. Having reached 80% two-dose coverage in 16+ year olds on 16 October, NSW crossed 90% coverage three weeks later on 8 November. At the same times, WA had only 55.6% and 67.4% coverage. Australia’s national coverage reached 80% on 5 November [38], and crossed 88% one month later on 5 December [39], days after the first community cases of Omicron were detected [40]. Our results indicate a substantial impact of increasing vaccination coverage from 70% to 80% and 90%. Considering the rate of the vaccine rollout that eventuated in Australian jurisdictions, the increase from 70% to 80% and to 90% coverage would have occurred in the first few weeks of the simulations had a dynamic rollout been implemented. As described in the results, we would anticipate PHSMs to change during different stages of an outbreak, particularly over the timescale that we have simulated. The 500-day time horizon was chosen to ensure that rapidly increasing outbreaks had resolved. We would not anticipate to see the outcomes presented here as, in addition to PHSMs changing, it is highly likely that new SARS-CoV-2 variants with varying characteristics would appear over this timescale [41].

We assumed in this analysis that infected individuals recover with complete protection from re-infection. At the time of these analyses, limited evidence was available on waning of immunity, particularly that induced by vaccination. A study of healthcare workers in England showed that reinfection was possible, with an estimated median interval between infections more than 200 days [15]. As this duration was of a similar magnitude to our simulated model scenarios, we did not expect waning immunity over this timeframe to alter our conclusions about relative differences between arrival pathways and vaccination thresholds. The reduced impact of vaccination against infection with the Omicron variant, however, would necessitate further analyses.

We made several assumptions about the quarantine pathways for the purpose of this work. First, we considered two settings for the volume of arriving travellers following reopening equivalent to 40% and 80% of pre-COVID volumes. These values were used for all quarantine pathways and durations, despite the fact that each would have different resourcing requirements. While this enabled straightforward comparison of various scenarios, a more nuanced analysis could also consider the resources required to facilitate quarantine of arriving travellers, particularly in the context of hotel quarantine. Similarly, we assumed that all arrivals were in groups of size four, and that the proportion of arrivals that were families was consistent with pre-COVID data. While it is likely that true arrival numbers and compositions differed from these simple assumptions, we anticipate little impact on the relative risks of different arrival pathways. Furthermore, Australia is a culturally diverse nation, with many multi-generational immigrants retaining close ties to their country of origin. We thus anticipated that many early arrivals (upon reopening) would be families disconnected for long periods by the border closures. Greater availability of data on arrival numbers and compositions would assist with more accurate estimation of absolute risks. Further, we implemented a fixed testing schedule within each quarantine pathway (days 1 and 5 for 7-day quarantine, days 1, 5 and 13 for 14-day quarantine), in line with national recommendations at the time [5]. We did not explore the impact of additional tests, their timing, or alternate turnaround times, as explored in, e.g., Wells *et al.* [42]. Wells *et al.* identified that testing on exit led to the lowest probability of onward transmission, with only a modest reduction following the addition of a test on entry. While this suggests that additional tests beyond those implemented in our analyses may not have a substantial impact, a key difference in our analyses to Wells *et al.* is the explicit risk posed by quarantine workers. Additional earlier tests may reduce the risk posed by quarantined infected arrivals to workers, though further work beyond the scope of this project would be required to quantify the impact this may have. Such analyses would need to consider the relative benefit of these additional enforced tests against the increased demand on laboratories, that may well be at capacity depending on the stage of an outbreak.

Finally, we assumed that all arriving travellers follow a single arrival pathway, and all arrivals came from settings with a pre-vaccination prevalence of 1% (0.2% with vaccination). The source prevalence assumptions resulted from the inability to reliably estimate the prevalence of infection in traveller countries of origin, given markedly disparate surveillance efforts and reporting. Given the relationship between the source prevalence and arrival volumes (equation (1)), our sensitivity analyses that consider double the arrivals can be equivalently considered as a doubling of the source prevalence. The assumption that arrivals enter via a single pathway was made with stakeholders to aid with interpretation of the relative impacts of the different quarantine pathways. A pragmatic alternative might involve filtering arrivals into different quarantine pathways according to their pre-arrival risk. For example, arrivals who test negative upon departure/arrival from a country with a high prevalence and/or low vaccine coverage (high-risk) may be directed to a longer duration or more stringent quarantine, while arrivals from a highly vaccinated country with low prevalence and high-testing coverage (low-risk) could quarantine for a shorter duration or at home. Different numbers of arrivals could then be filtered through each pathway accordingly, increasing the number of arrivals that could be sustained. While only individual pathways are reported here, the model framework is readily able to support more complex combinations of arrival pathways— the linking model code permits a user to specify a number of arrivals and source prevalence arriving into each of the arrival pathways (quarantine type and duration). Arrivals linelists that combine arrival volumes and pathways are then generated using our linking model, and feed into the community transmission model. To implement this, the number of arrivals that present through each of the arrival pathways for standard and family arrival groups are entered into the columns of a data frame, num.arrivals.mat, within the linking model. Each row of this data frame represents a different scenario, combining arrival volumes through each pathway and generating the arrival linelist. The desired source prevalence associated with the arrivals into each pathway is specified in the corresponding entries of another data frame, path.prev.mat. By structuring the model in this way, our framework can generate timely risk assessments in the event of future pandemics.

### Summary

While effective at limiting importation of infection, border controls and quarantine measures can have a profound social [43] and economic [44] impact on populations. Modelling is therefore critical to ensure that such measures are proportionate to the benefit they provide. Reopening international borders during a pandemic when infection is still circulating globally requires careful evaluation and management of importation and outbreak risks. We have demonstrated how a model-based analysis can be used to assess how importation and outbreak risks vary with travel volume, arrival and quarantine pathways, and the epidemiological characteristics of the community.

## Data Availability

No new datasets are presented in this research. Code to perform the analyses is available at

https://github.com/aus-covid-modelling/NationalCabinetModelling-OctNov21

## Acknowledgements

We would like to thank Nancy Shi for helping to generate the workflow schematic. Computations were supported by use of the Nectar Research Cloud, a collaborative Australian research platform supported by the National Collaborative Research Infrastructure Strategy (NCRIS); MASSIVE HPC facility (www.massive.org.au); and The University of Melbourne’s Research Computing Services.

## Author Contributions

M.J.L.: conceptualisation, methodology, software, formal analysis, writing–review & editing; C.Z.: conceptualisation, methodology, writing–review & editing; E.C.: methodology, writing - review & editing; F.M.S.: conceptualisation, methodology, writing–review & editing; C.M.B.: conceptualisation, writing–review & editing; J.V.R.: conceptualisation, writing–review & editing; J.C.M.: conceptualisation, writing–review & editing; J.M.M.: conceptualisation, writing–review & editing; N.G.: conceptualisation, methodology, writing–review & editing, supervision; J.M.: conceptualisation, writing–original draft, writing–review & editing, supervision; D.J.P.: conceptualisation, methodology, formal analysis, writing–original draft, writing–review & editing, visualisation, supervision.

