## Supplementary material for "Border quarantine, vaccination and public health measures to mitigate the impact of COVID-19 importations in Australia: a modelling study"

### S1 Supplementary Methods

The following provides results showing the vaccination coverage and distribution of time from infection to isolation, as referenced in Methods.

In late August 2021, the Australian Technical Advisory Group for Immunisation (ATAGI) recommended vaccination for 12–15 year olds. At the time this work was completed in late-October 2021, 16–29 year olds had just been made eligible for vaccination within the age-based vaccination rollout (from 11 October 2021). Further, vaccination was not approved for individuals under 12 years of age, and so the coverage in that group is assumed to be zero. The ChAdOx1 vaccine was restricted for use only in individuals aged over 60 early in Australia's vaccination rollout, leading to high proportions of coverage with that vaccine in older individuals, while the Comirnaty vaccine was used in younger individuals. The Spikevax vaccine was not used until very late in Australia's vaccine rollout, hence the low prevalence of that vaccine in the population. The coverage values in Figure S1 were provided by the model-based estimates of the vaccine rollout, described in Methods and [1].

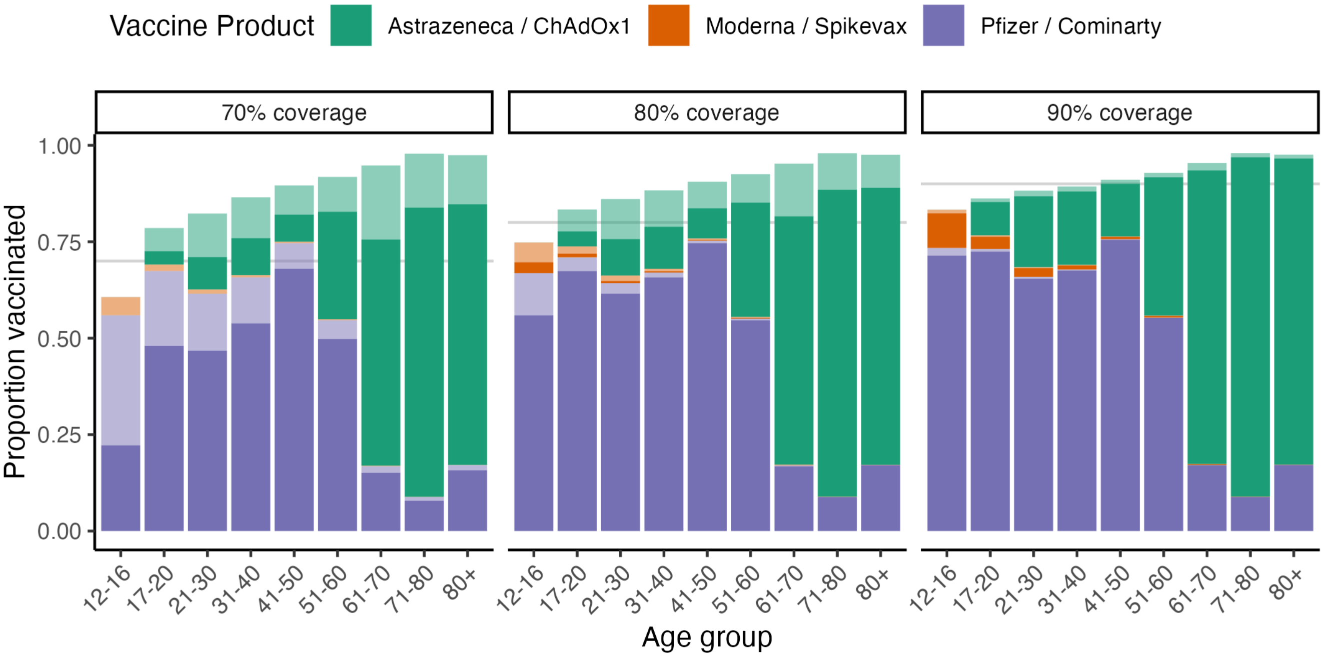


**Fig S1.** Modelled one- (light colour) and two-dose (dark colour) two-dose coverage by age group and vaccine product, at 70%, 80% and 90% vaccination coverage. Used to define the community characteristics in the community model at the specified vaccination coverage.

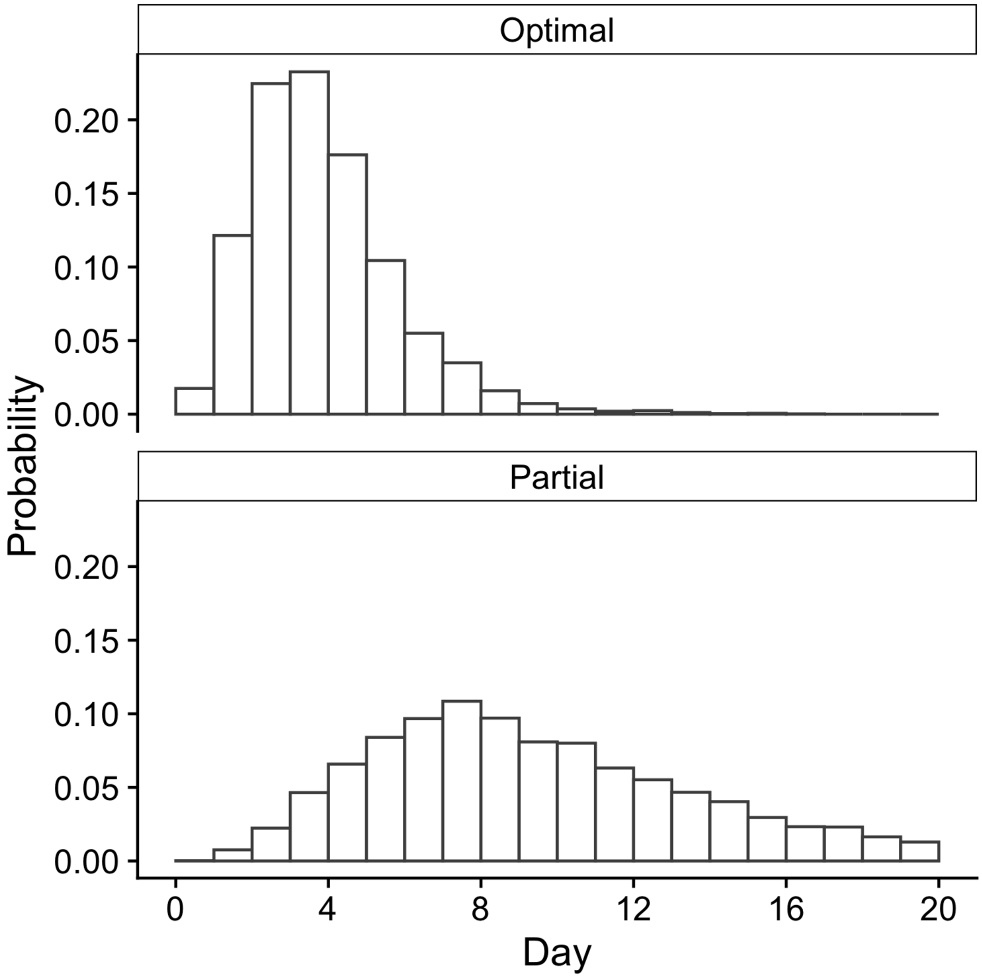


**Fig S2.** Distribution of time from infection to isolation. These distributions are used to incorporate the effects of the test-trace-isolate-quarantine systems in the community, by modifying an individual's probability of transmission at a given time. Further description provided in [2]

#### S1.1 Quarantine model: within-host model

The quarantine model captures individual-level variability in the time-varying level of infectiousness and test sensitivity, representing the progression of the pathogen within a person. Full details of these model components can be found in [3] and [4], based on [5], but are briefly presented below.

##### S1.1.1 Time-varying infectiousness

We define a piecewise function that describes an individual's infectiousness as: i) increasing from infection throughout their incubation period, ii) plateauing at a peak level of infectiousness from a time prior to symptom onset (irrespective of whether the individual is symptomatic or asymptomatic), and iii) declining following symptom onset to a recovery time.

In particular, we use the following equations to describe infectiousness over time since infection for individual $j$:

$$\beta_j(t) = \frac{z_j(t) - z_j^{min}}{z_j^{max} - z_j^{min}} z_j^{max}, \tag{1}$$

where,

$$z_j(t) = \begin{cases} z_j^{max}\exp(k_j^{inc}t)V_{max}^{-1} & \text{for } \tau_0 < t \leq \tau_1 \\ z_j^{max} & \text{for } \tau_1 < t \leq \tau_2 \\ z_j^{max}\exp(k_j^{rec}t) & \text{for } \tau_2 < t \leq \tau_{rec} \end{cases} \tag{2}$$

where, for individual $j$'s incubation period duration $t_j^{\text{inc}}$, and the duration of the peak infectiousness $t_j^{\text{peak}}$,

- $\tau_0$ corresponds to the time of infection
- $\tau_1 = \tau_0 + t_j^{\text{inc}} - t_j^{\text{peak}}$,
- $\tau_2 = \tau_1 + t_j^{\text{peak}}$, and
- $\tau_{\text{rec}} = \tau_2 + t_j^{\text{rec}}$.

All parameters describing the individuals time-varying infectiousness are defined in Table S1 below.

**Table S1.** Individual-level parameters governing an infected person's trajectory of infectiousness over time.

| Parameter | Value | Description |
|---|---|---|
| $z_j^{\text{max}}$ | $z^{\text{max}} \sim \text{Gamma}(\phi = 0.15, \langle\beta\rangle\phi^{-1})$ | peak (maximum) infectiousness |
| $\langle\beta\rangle$ | $1.52\ (\frac{R_0}{3.94})$ | mean peak infectiousness ($R_0 = 6$, see [3] for calibration details) |
| $t_j^{\text{inc}}$ | $t^{\text{inc}} \sim \text{Log-normal}(\mu = 1.62, \sigma = 0.418)$ | incubation period between infection and symptom onset (mean = 5.51 days) |
| $t_j^{\text{peak}}$ | $t_j^{\text{inc}}\Delta_{\text{peak}}$ | duration of infectiousness peak |
| $\Delta_{\text{peak}}$ | 0.1 | fraction of incubation period over which infectiousness plateau lasts |
| $t_j^{\text{rec}}$ | $t^{\text{rec}} \sim \text{Uniform}(5, 10)$ | period between symptom onset and recovery |
| $k_j^{\text{inc}}$ | $\log(V_{\text{max}})[t_j^{\text{inc}} - t_j^{\text{peak}}]$ | growth rate of infectiousness during incubation |
| $k_j^{\text{rec}}$ | $\log([V_{\text{max}}]^{-1})[t_j^{\text{rec}}]^{-1}$ | decay rate of infectiousness during recovery (e.g., after onset of symptoms) |
| $V_{\text{max}}$ | 7.0 | shape parameter controlling steepness of growth and decline of infectiousness |

##### S1.1.2 Time-varying test sensitivity

Similar to the time-varying infectiousness, test-sensitivity varies over time relative to the time since infection and symptom onset. The functional form and parameters were based on results from [5]. A piecewise function describes the increase in test-sensitivity following infection up to a time prior to symptom onset (irrespective of whether the individual is symptomatic or asymptomatic), and the decline after symptom onset. The probability of an individual testing positive given they are infected is given by:

$$Pr(\text{test positive at time } t \mid \text{infected}) = \begin{cases} [1+\exp(-(b_1+b_2t))]^{-1} & t \le T_c \\ [1+\exp(-(b_1+b_2t+b_2b_3t))]^{-1} & t > T_c \end{cases} \quad (3)$$

where $T_c$ is a time sampled for each individual $j$ relative to their symptom onset $t_j^{inc}$,

$$T_{c,j} = t_j^{inc} - 4.11q_j \quad (4)$$

where $q_j$ is the quantile corresponding to the sampled incubation period (i.e., the CDF at the sampled incubation period for individual $j$). A negative correlation between an individuals incubation period and the rate test sensitivity increases prior to $T_c$ was imposed such that,

$$b_{2,j} = 1.26 - 2.11(1 - q_j). \quad (5)$$

Parameters $b_1$ and $b_3$ are sampled independently from uniform distributions, $U(0.8, 2.31)$ and $U(-1.14, -1.05)$, respectively.

#### S1.2 Quarantine Pathways

In this study we compare two different quarantine pathways, one in which arriving travellers stay within dedicated quarantine hotels, and another in which they quarantine within private dwellings. These two pathways both implement screening and case isolation, subject to the constraint that children remain in the company of at least one adult. This section provides additional details regarding the structural features of the different quarantine pathways, screening strategies, and response to case detection.

**Screening and Isolation**

Travellers arriving into quarantine are assumed to be either uninfected, pre-symptomatic, or asymptomatic, and arrive in groups of four close contacts. After arrival into quarantine, travellers are tested on days 1, 5, and 13 (14-day quarantine) or on days 1 and 5 (7-day quarantine). If a traveller tests positive or presents symptomatic illness, they are put into case isolation for a period of 10 days, and subsequently released from quarantine. The quarantine period for their close contacts is extended by 14 days – regardless of whether they initially entered a 7- or 14-day quarantine – and the testing schedule is reset (test on days 1, 5, and 13 of the extension period). If a traveller in extended quarantine tests positive or presents symptoms, they are subject to 10-days of case isolation, but the quarantine period of their close contacts is not extended further. The testing, case isolation, and quarantine extension response rules are depicted in Figure S3.

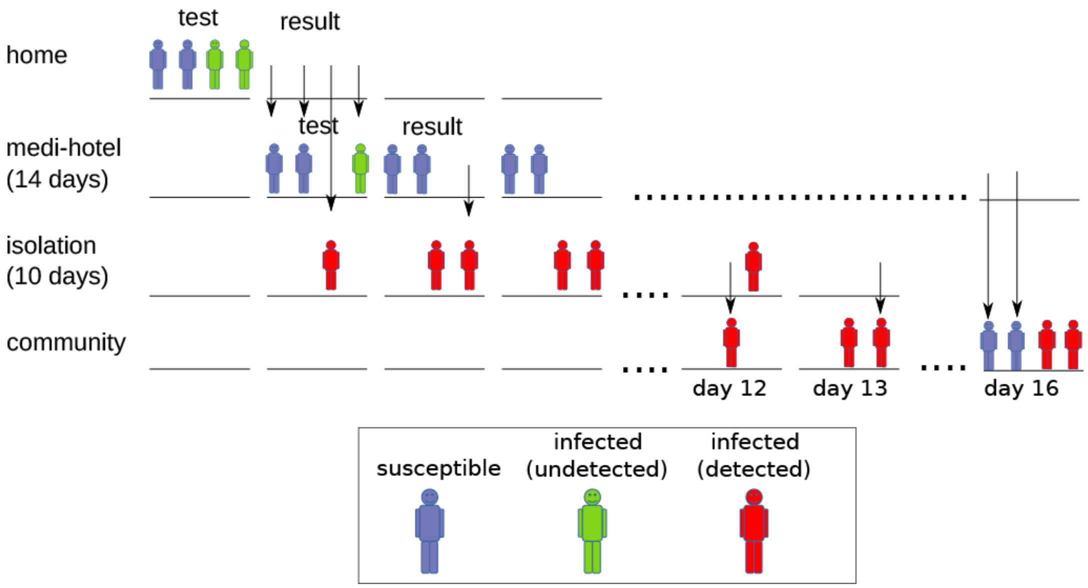


**Fig S3.** Schematic of the infection screening and response process for a single group of travellers moving through home quarantine. Case detection triggers a 10-day isolation period for travellers who test positive. Detection also triggers the transfer of close contacts from the home into a "medi-hotel" for a period of 14 days. Travellers are allowed to enter the community after these extended quarantine periods end, regardless of infection status (release does not require a negative test).

##### Family Groups in Quarantine Case Isolation

When simulating family units in quarantine, travelling groups of 4 close contacts are assumed to contain two adults and two children. The strategy for splitting family groups to isolate known cases is shown in Figure S4. Depending on which members of the group test positive (or present symptoms), the group is split so as to minimise the potential for other members to become infected, while also ensuring that each child remains in the company of an adult.

##### Hotel Quarantine

Structurally, the hotel quarantine system has three compartments: the quarantine hotel compartment, the "medi-hotel" compartment, and the case isolation compartment. In the hotel compartment, there is potential for transmission between different groups of close contacts, and between travellers and hotel workers. In the "medi-hotel", transmission may only occur between close contact groups, and in case isolation, no transmission is possible.

In the hotel quarantine compartment, infectious contacts between travellers in different groups are subject to reduced force of infection (by a factor of 0.01 relative to same-group contacts), with the same factor applied to contacts between travellers and workers. On the other hand, infectious contacts between workers are reduced by a factor of 0.1 relative to unmitigated contact between travellers in the same group, to simulate the reduction in transmission potential from judicious use of personal protective equipment by workers. A schematic of the hotel quarantine pathway is shown in Figure S5.

##### Home Quarantine

The home quarantine pathway is similar to that of hotel quarantine, with two main differences. The first is that contacts are not allowed between different groups of travellers (and no workers are present in the home environment). The second key difference is that, due to imperfect compliance, travellers in quarantine have intermittent contact with the outside community. The amount of contact is determined by the *compliance* parameter which takes a value between 0 and 1, and is equivalent to the probability that an individual will not make contact with members of the outside community on a given day. A schematic of the home quarantine pathway is shown in Figure S6.

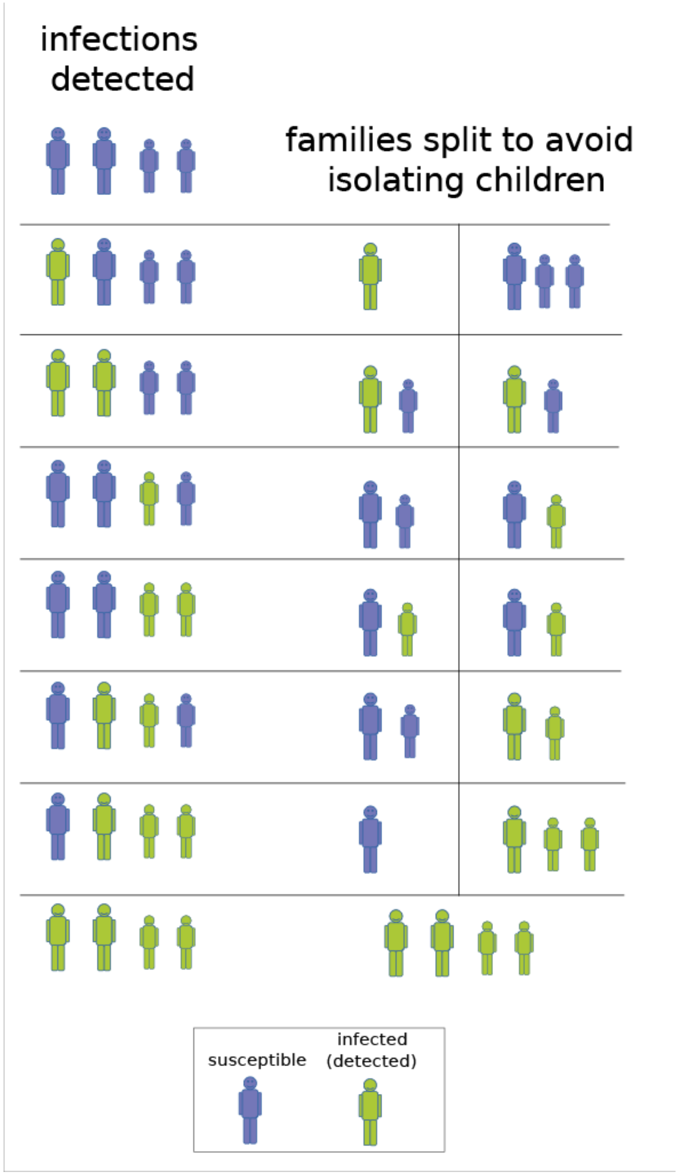


**Fig S4.** Schematic of case isolation strategies for family groups containing children accompanied by adults. The "infections detected" column illustrates the potential combinations of infected individuals within a family group (children are represented as smaller in size). For each configuration of infected individuals, the second column illustrates how the group is split in order to minimise transmission potential while ensuring children remain in the company of an adult during case isolation.

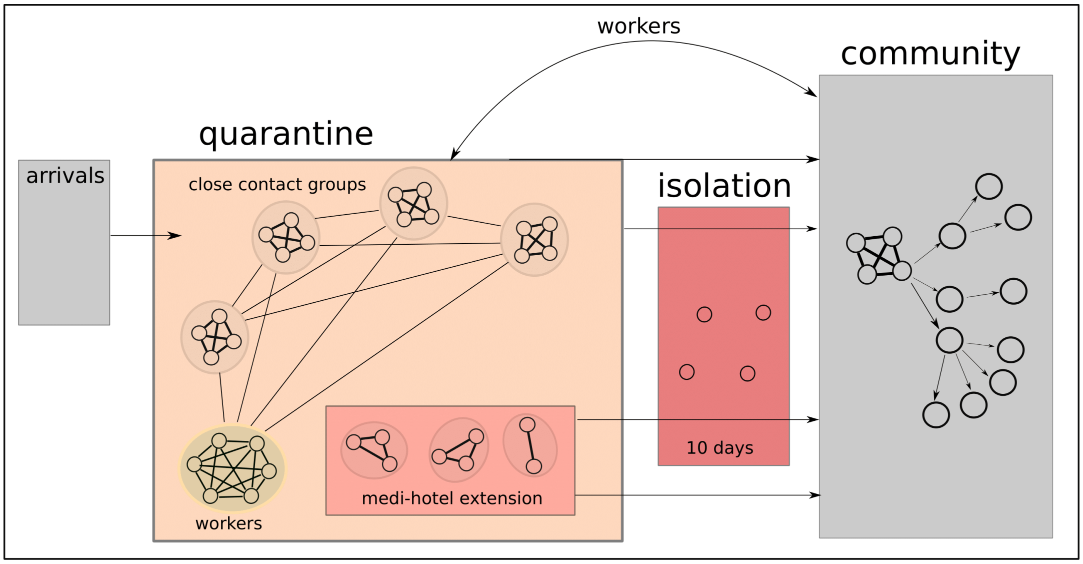


**Fig S5.** Schematic of hotel quarantine for arriving travellers. After arrival, groups of close contacts are housed in hotel rooms. Contact between travellers in the same groups is unmitigated, while contact rates between groups of travellers is reduced. The quarantined groups are also in contact with the hotel workforce, with reduced transmission potential. Detected infections are placed into isolation, with their close contacts placed into a “medi-hotel” extension which eliminates transmission potential outside of close contact groups. After the quarantine period ends, travellers enter the community where any who remain infected may generate outbreaks of community transmission. While in hotel quarantine, infected travellers may contribute to community transmission by infecting quarantine workers who maintain continued community interactions.

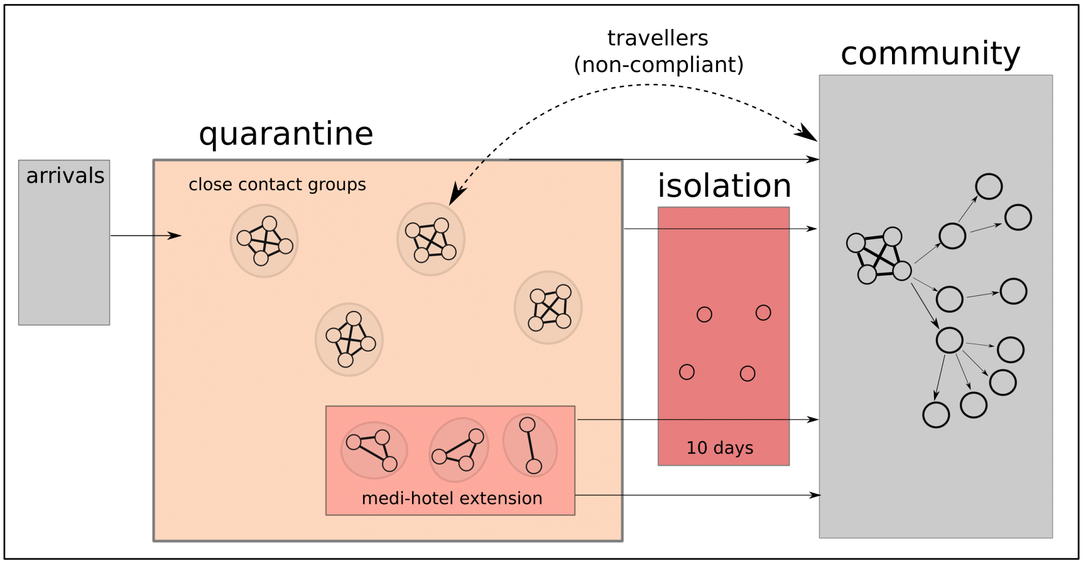


**Fig S6.** Schematic of home quarantine for arriving travellers. After arrival, travellers remain in private dwellings during their quarantine period. While case response is handled in the same manner as for hotel quarantine (case isolation and extension for contacts), travellers quarantining at home have intermittent direct contact with the community due to sporadic non-compliance.

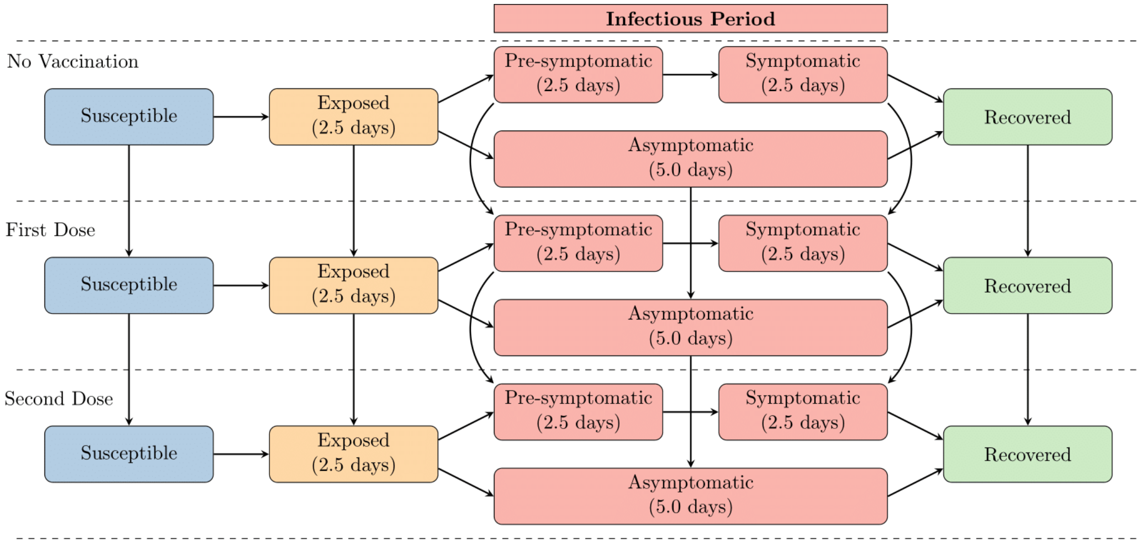


**Fig S7.** Simplified schematic of community transmission model reproduced from [1]. For the purposes of this work, the force of infection was modified by the introduction of a quarantine breach event into this population, with the community transmission dynamics then following as described above. Vaccine coverage at one- and two-dose was fixed (i.e., no transition from no vaccination to first or second dose) according to the vaccination coverage thresholds specified, shown in Supplementary Figure S1.

#### S1.3 Community transmission model

Vaccine rollout was not explicitly simulated over time in the community transmission model for this work (as in the model presented in Conway *et al.* [1]), but implemented as one of several achieved threshold coverage levels in the age-eligible population, shown in Figure S1. Individuals from each age bracket are vaccinated according to these coverage thresholds via uniform random sampling, and their vaccine status recorded in the model.

The infection model is based on the Susceptible-Exposed-Infected-Recovered (SEIR) paradigm. A simplified diagram of the infection model is provided in Figure S7. Briefly, the individuals in our simulation are susceptible until they are infected by an infectious contact, at which point they will transition into the exposed state. At a later time, the exposed individual will move into the infectious state and be pre-symptomatic. This is the period where the individual is infectious but is currently not displaying any symptoms. After a finite amount of time in the pre-symptomatic state, the individual will become either asymptomatic or symptomatic. Finally, the individual will transition into the recovered state and no longer be infectious. Note that for our purposes the actual dynamics within each compartment are more complicated, depending upon all state variables of the individuals, the specifics of which will be detailed here.

To simulate infection within the model, we explicitly track the exposed and infected individuals in two dynamic arrays. It is only important to track the exposed and infected individuals as those that are susceptible and recovered do not directly contribute to the spread of SARS-CoV-2. At each time-step we update all individuals in the exposed dynamic array, followed by updating all individuals in the infected dynamic array, tracking all transitions between compartments for the next time-step. For each of the individuals in the exposed dynamic array we determine whether or not they will become infectious at this point in time. This is done by checking against the member variable that contains the time they become infectious. If the individual becomes infectious at this time-step, they are removed from the exposed dynamic array and added to the end of the infected array for the next time-step (care must be taken to ensure that the newly infectious individual is not updated twice in a single time-step now that they are in the infected array).

The individuals in the infected dynamic array are updated according to the contacts, transmission, and vaccine-induced protection as described in the 'Community transmission model' in the main text Methods. Parameters relating to these processes are presented in Supplementary Table S2.

The exposure of a contact sets the appropriate state variables of the newly infected individual. To do so, we set their infection status to exposed and add their information to the dynamic array of exposed individuals. At this point, we set the individual's time of exposure and sample the time that the individual will transition to the infectious state, the time that they will develop symptoms, the time of isolation given symptom onset and the time that they will recover. Using the known vaccine status of the newly exposed individual, we determine if they will develop symptoms and assign their transmissibility for the entire infectious period.

To determine if the individual is asymptomatic or symptomatic we use a Bernoulli trial with probability $q_i = (1 - VE_q)q_i^0$, where $VE_q$ is the vaccine efficacy against symptoms and $q_i^0$ is the baseline probability of symptoms for an individual in age bracket $i$. The transmissibility of the newly infected individual is defined at exposure and is assumed to remain constant throughout the individual's entire infectious period. Once all contacts and infection events have been simulated for the infectious individual, we check if they either become symptomatic or recover.

To determine if an individual becomes symptomatic, we check if the symptom timer should be triggered. To determine if the individual recovers we check if the recovery timer has triggered. At the time of recovery, the infectious individual's infection status is changed to recovered and they are removed from the dynamic array that tracks infectious individuals. It is assumed that once recovered, the individual will remain recovered for the rest of the simulation (no waning immunity). Note that due to the assumption of no waning immunity, we do not have to be vigilant in resetting any state variables within the individual. As they are recovered, we ensure that all further infection events in which they are involved fail. Finally, we store all statistics of the infection event for output at the end of the simulation.

**Table S2.** Key vaccination, transmission and simulation parameters from the community transmission model.

| Parameter | Value | Reference |
|---|---|---|
| **Relative protection ($\tau$)** | (Dose 1, Dose 2) | |
| Pfizer/Moderna | (0.914, 0.164) | [6,7] |
| Astrazeneca | (1.09, 0.519) | [6,7] |
| **Transmission Potential** | | |
| NSW | 4.75 | [2] |
| WA | 6.32 | |
| **% arrivals families** | | |
| NSW | 17.9% | Provided by |
| WA | 18.2% | government |
| Initial Infection | 200 | |
| Simulation timestep | 1 day | |
| $t_{\max}$ | 500 | |
| Infectious period | | |
| Presymptomatic period | $\Gamma(1, 1.5)$ | [8] |
| Symptomatic period | $\Gamma(1.01, 4.5)$ | [9] |
| Exposed period | $\Gamma(4.82, 0.52)$ | [10] |
| Reduced infectiousness for children | 0.6 | [11,12] |

### S2 Supplementary Results

#### Setting with no existing transmission

Figures S8 and S9, and Tables S3 and S4 report on the relative infections under each quarantine and vaccination setting, respectively, for the setting with no existing transmission, low PHSMs and optimal TTIQ.

**Table S3.** The infections attributable to imported cases for each quarantine pathway relative to the average number with no quarantine in place (bottom row). Setting with no transmission, and baseline PHSMs in place is shown, corresponding to Figure S8. Note that these values are indicative only, as not all epidemics have not resolved over the 500 day time horizon. Values are presented as the mean with $(25,75)^{th}$ percentiles.

| Quarantine | Total (Imported) |
|---|---|
| 7-day Home | 53.85 (30.42, 76.73) |
| 14-day Home | 19.59 (0.46, 30.87) |
| 7-day Hotel | 37.99 (97.04, 67.64) |
| 14-day Hotel | 6.17 (0.01, 0.03) |
| 14-day Hotel (unvaccinated) | 39.41 (8.14, 67.33) |
| No quarantine | 100.00 (92.89, 109.99) |

**Table S4.** The infections attributable to imported or local sources, and overall, for each vaccination coverage relative to the average number with 70% vaccination coverage. Setting with existing transmission for 7-day Home quarantine, with baseline or low PHSMs in place, corresponding to Figure S9. Note that these values are indicative only, as not all epidemics have not resolved over the 500 day time horizon. Values are presented as the mean with $(25,75)^{th}$ percentiles.

| Vaccination Coverage | Total (Imported) |
|---|---|
| 90% | 2.17 (1.06, 3.01) |
| 80% | 19.74 (14.82, 25.41) |
| 70% | 100.00 (99.13, 100.90) |

#### Setting with existing transmission

Figures S10 and S11–S13, and Tables S5 and S7 report on the relative infections under each quarantine and vaccination setting, respectively, for the setting with no existing transmission, baseline/low PHSMs and partial TTIQ.

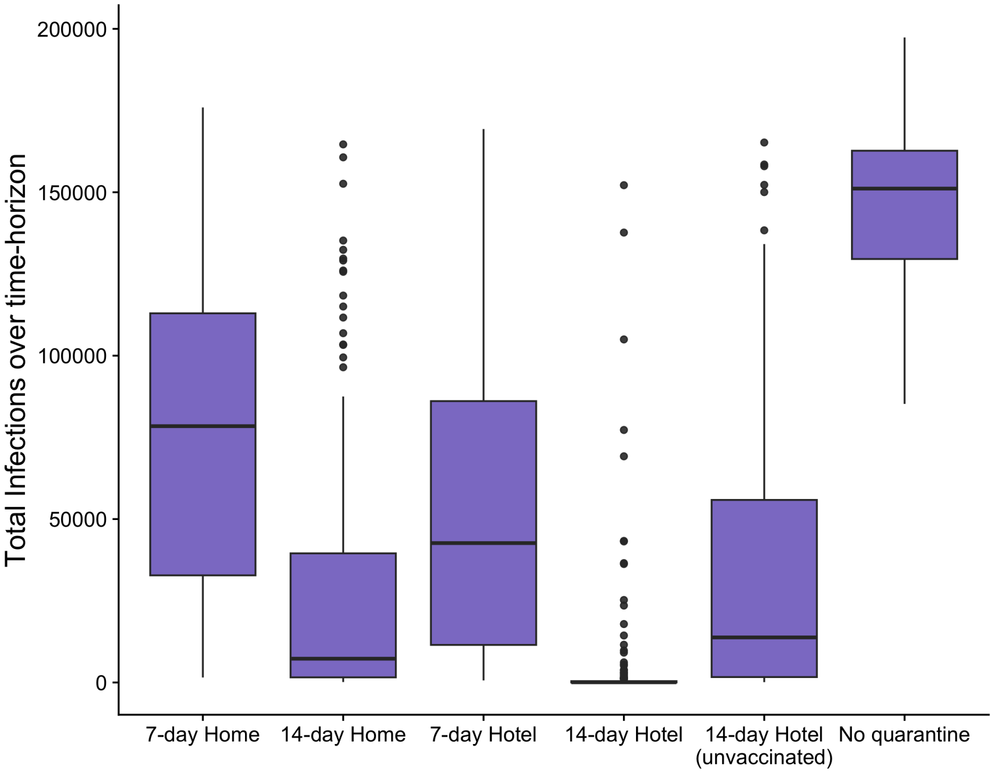


**Fig S8.** Box plots of the relative number of infections for each quarantine strategy in the setting with existing transmission and baseline PHSMs. Number of infections are relative to the average number observed under the 'No quarantine' scenario, indicated by the horizontal dashed line.

**Table S5.** The infections attributable to imported or local sources, and overall, for each quarantine pathway relative to the average number with no quarantine in place (bottom row). Setting with existing transmission, and baseline PHSMs in place is shown, corresponding to Figure S10. Values are presented as the mean with $(25,75)^{th}$ percentiles.

| Quarantine | Imported | Local | Total |
|---|---:|---:|---:|
| 7-day Home | 5.21 (2.22, 7.33) | 93.06 (91.05, 96.17) | 98.26 (97.29, 99.29) |
| 14-day Home | 1.46 (0.26, 1.91) | 96.47 (95.37, 97.95) | 97.93 (97.06, 98.82) |
| 7-day Hotel | 2.84 (0.90, 3.44) | 95.18 (93.91, 97.30) | 98.02 (97.04, 99.17) |
| 14-day Hotel | 0.37 (0.02, 0.17) | 97.24 (96.39, 98.38) | 97.61 (96.69, 98.61) |
| 14-day Hotel (unvaccinated) | 3.35 (1.25, 4.05) | 94.77 (93.59, 97.08) | 98.12 (97.20, 98.94) |
| No quarantine | 21.40 (14.50, 25.28) | 78.60 (74.08, 85.53) | 100.00 (99.12, 100.82) |

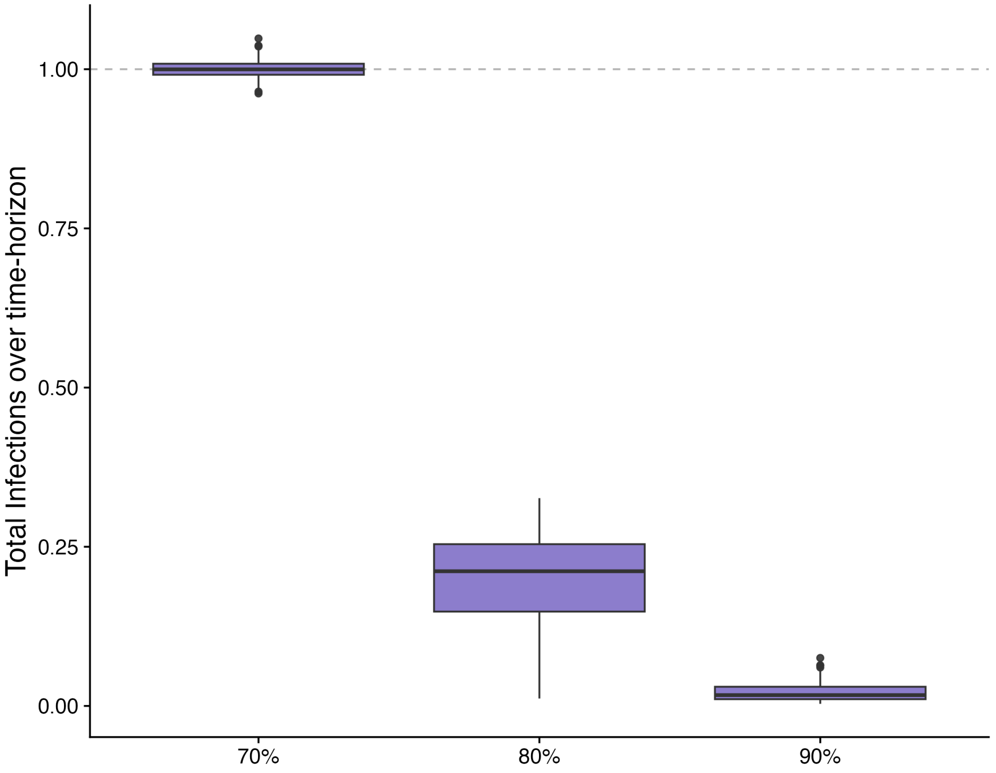


**Fig S9.** Box plots of the relative number of infections for each quarantine strategy in the setting with existing transmission and baseline PHSMs. Number of infections are relative to the average number observed under the 'No quarantine' scenario, indicated by the horizontal dashed line.

**Table S6.** The infections attributable to imported or local sources, and overall, for each quarantine pathway relative to the average number with no quarantine in place (bottom row). Setting with existing transmission, and low PHSMs in place is shown, corresponding to Figure S10. Values are presented as the mean with $(25,75)^{th}$ percentiles.

| Quarantine | Imported | Local | Total |
|---|---|---|---|
| 7-day Home | 19.28 (15.33, 21.60) | 7.15 (4.63, 8.67) | 26.43 (22.02, 30.00) |
| 14-day Home | 5.34 (3.30, 6.52) | 7.29 (4.83, 8.71) | 12.63 (9.19, 15.24) |
| 7-day Hotel | 12.77 (9.34, 16.04) | 7.12 (4.71, 8.43) | 19.89 (15.63, 23.82) |
| 14-day Hotel | 1.63 (0.44, 2.08) | 7.67 (4.83, 9.57) | 9.30 (6.61, 11.23) |
| 14-day Hotel (unvaccinated) | 14.00 (10.22, 17.31) | 6.95 (4.361, 8.41) | 20.95 (16.36, 24.76) |
| No quarantine | 92.80 (85.31, 100.29) | 7.20 (4.571, 8.75) | 100.00 (91.92, 108.38) |

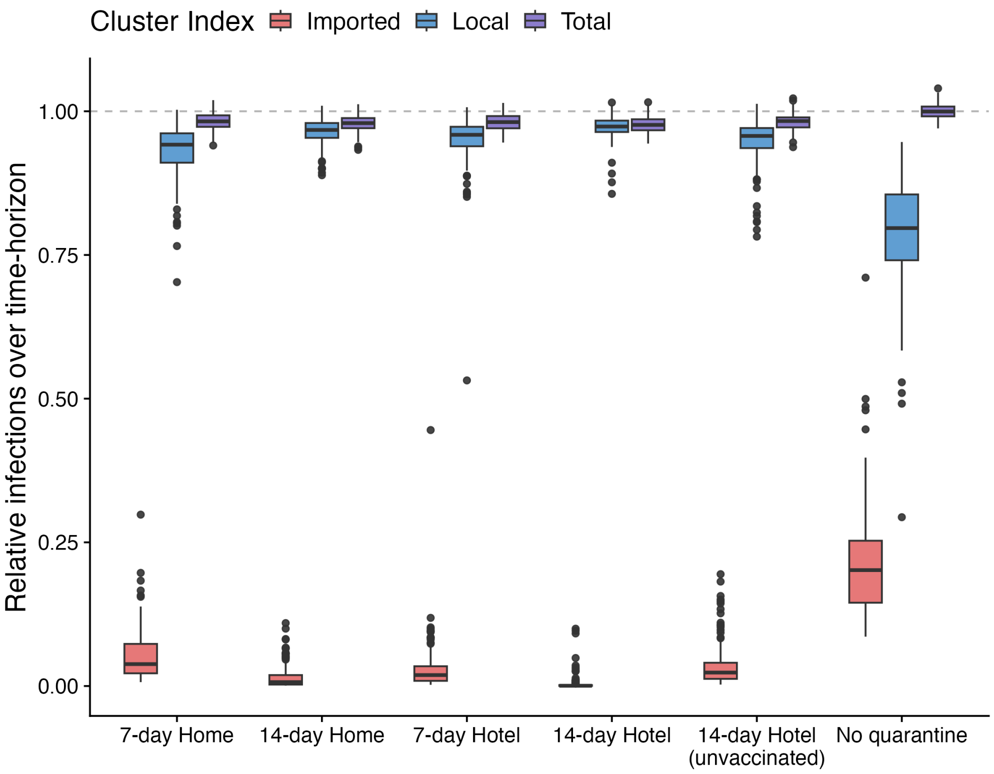


**Fig S10.** Box plots of the relative number of infections (purple), and those generated through transmission chains initiated by an arrival (red) or a local case (blue) for each quarantine strategy in the setting with existing transmission and baseline PHSMs. Number of infections are relative to the average number observed under the 'No quarantine' scenario, indicated by the horizontal dashed line.

**Table S7.** The infections attributable to imported or local sources, and overall, for each vaccination coverage relative to the average number with 70% vaccination coverage. Setting with existing transmission for 7-day Home quarantine, with baseline (corresponding to Figure S12) or low (corresponding to Figure S13) PHSMs in place. Values are presented as the mean with $(25,75)^{th}$ percentiles.

| PHSMs | Vaccination Coverage | Imported | Local | Total |
|---|---|---|---|---|
| Baseline | 90% | 6.01 (2.82, 7.76) | 17.73 (15.89, 21.03) | 23.74 (22.93, 24.73) |
| | 80% | 4.58 (2.16, 5.90) | 48.58 (47.30, 50.73) | 53.16 (52.54, 53.74) |
| | 70% | 2.32 (1.27, 2.92) | 97.68 (96.95, 98.76) | 100.00 (99.64, 100.4) |
| Low | 90% | 8.18 (7.15, 9.02) | 1.91 (1.38, 2.24) | 10.09 (8.88, 11.01) |
| | 80% | 13.86 (11.31, 15.95) | 3.25 (2.18, 4.05) | 17.11 (14.50, 19.24) |
| | 70% | 66.93 (47.94, 84.73) | 33.07 (10.93, 50.51) | 100.00 (74.38, 123.4) |

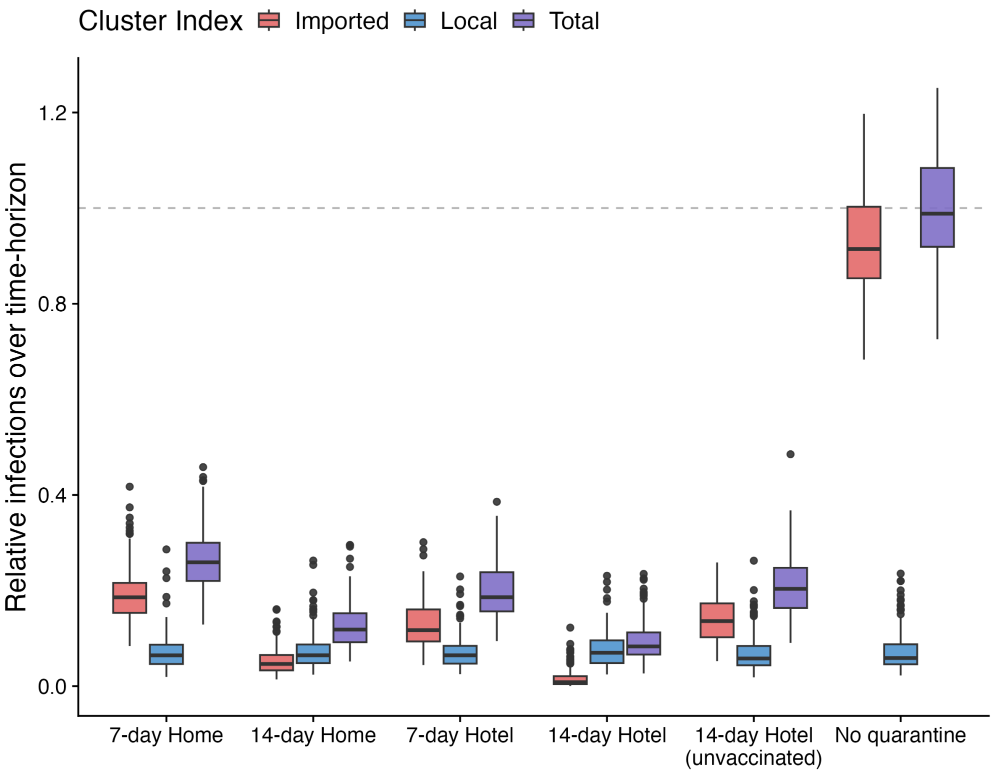


**Fig S11.** Box plots of the relative number of infections (purple), and those generated through transmission chains initiated by an arrival (red) or a local case (blue) for each quarantine strategy in the setting with existing transmission and low PHSMs. Number of infections are relative to the average number observed under the 'No quarantine' scenario, indicated by the horizontal dashed line.

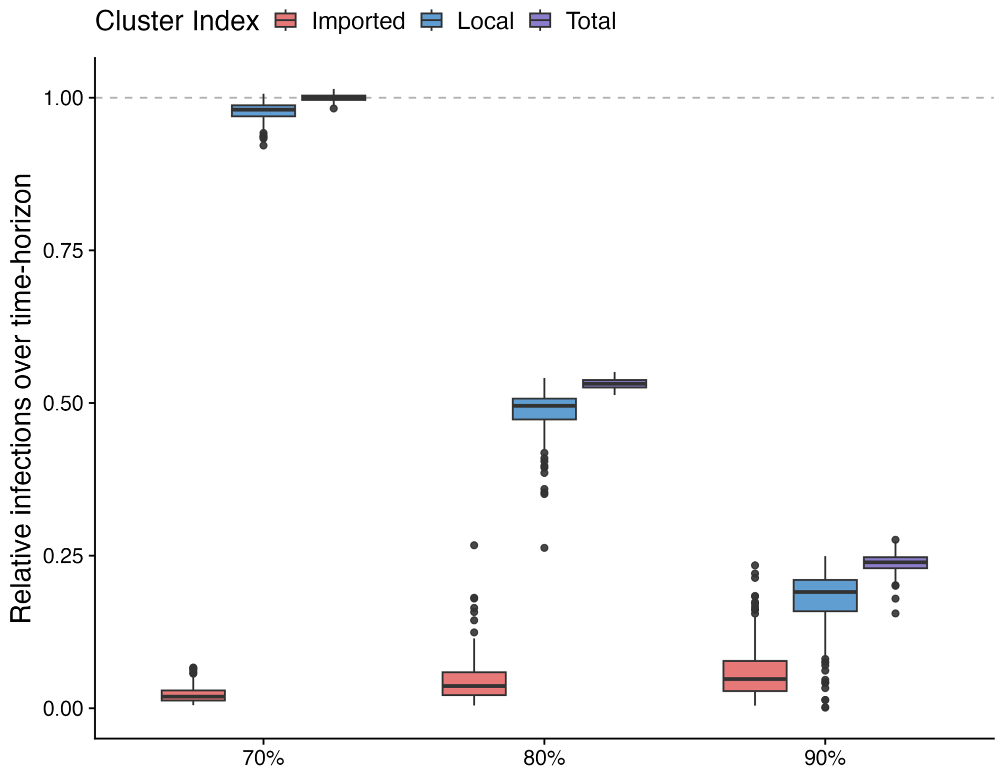


**Fig S12.** Box plots of the relative number of infections (purple), and those generated through transmission chains initiated by an arrival (red) or a local case (blue) for each quarantine strategy in the setting with existing transmission and baseline PHSMs. Number of infections are relative to the average number observed under the 'No quarantine' scenario, indicated by the horizontal dashed line.

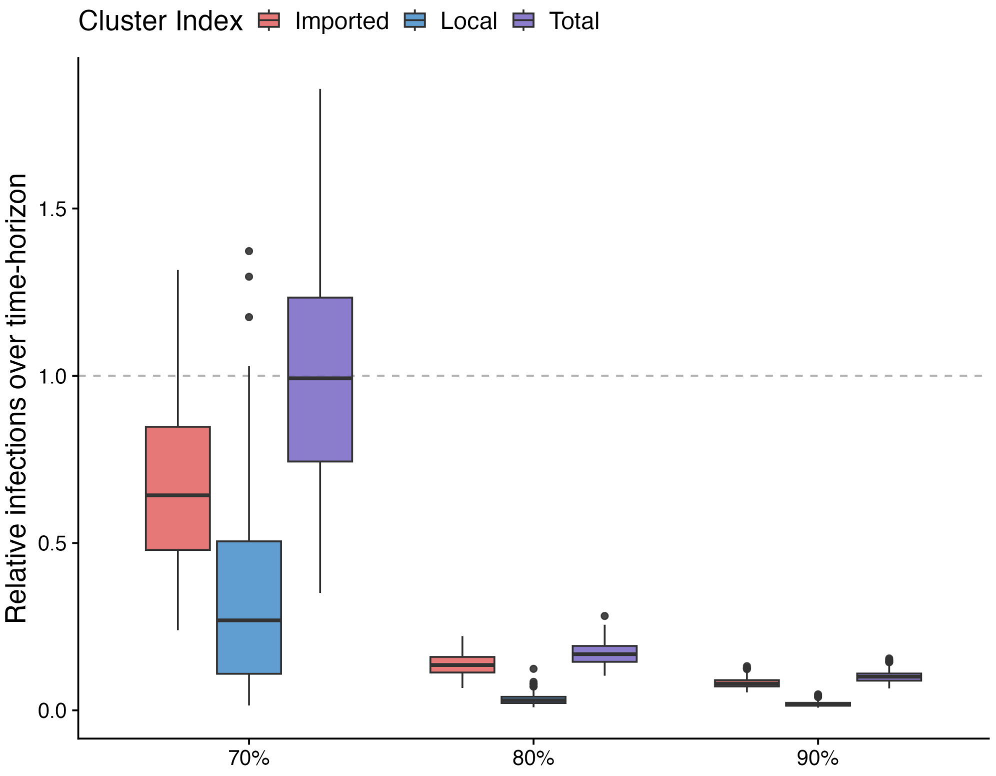


**Fig S13.** Box plots of the relative number of infections (purple), and those generated through transmission chains initiated by an arrival (red) or a local case (blue) for each quarantine strategy in the setting with existing transmission and baseline PHSMs. Number of infections are relative to the average number observed under the 'No quarantine' scenario, indicated by the horizontal dashed line.

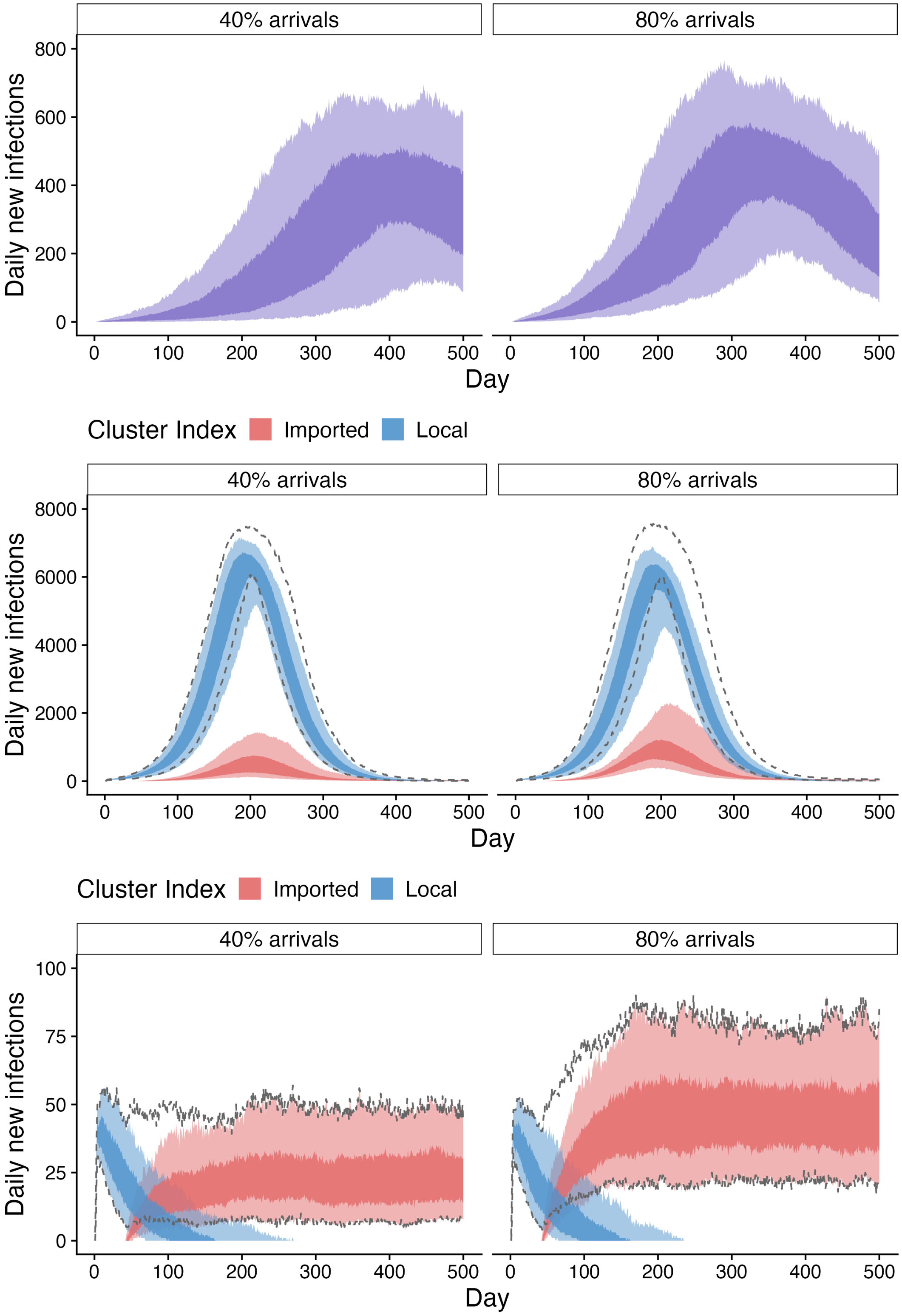


**Fig S14.** Daily new infections as a result of imported infections via 7-day home quarantine into a community with no existing transmission (top), or existing transmission with baseline (middle) or low (bottom) PHSMs, where arrival volumes are 40% (left) or 80% (right) of 2019 levels. Community vaccination is 80%. In the community with existing transmission, colour represents whether outbreak was seeded by a locally derived case, or an imported case. All infections in the community with no existing transmission are seeded by imported infections. Dark and light ribbons represent 50% and 90% intervals, respectively. Arrivals initiate on day 40.
